# Developing Meaningful Score Differences for the Bayley-4 and Vineland-3 in Angelman Syndrome using a Delphi Panel

**DOI:** 10.1101/2025.04.05.25325305

**Authors:** Sonya Powers, Katherine N. Anderson, Wen-Hann Tan, Angela Gwaltney, Sarah Nelson Potter, Julian Tillmann, Mark Daniel, Audrey Thum, Cristan Farmer, Susanne Clinch, Lisa Squassante, Jorrit Tjeertes, Brenda Vincenzi, Katalin Buzasi, Anne C. Wheeler, Anjali Sadhwani, the Meaningful Change Delphi panel

## Abstract

**Objectives:** To develop within-patient meaningful score differences (MSDs) on the Bayley Scales of Infant Development, Fourth Edition (Bayley-4), and the Vineland Adaptive Behavior Scales, Third Edition (Vineland-3), for individuals with Angelman syndrome (AS).

**Methods:** A Delphi method, involving a panel of 19 caregivers of individuals with AS, was used to establish MSDs for Bayley-4 and Vineland-3 Growth Scale Values. MSD was defined as the smallest change that would noticeably impact the daily functioning of an individual with AS or family quality of life in a way that was important to the individual with AS or their family. For each subscale of the Bayley-4 and Vineland-3, the panel was presented with 2 to 4 vignettes describing varying levels of baseline functioning and asked to select a MSD from a range of potential values. An iterative process involving three rounds of ratings and two rounds of discussion was used to build consensus. The median caregiver rating from round 3 was retained as the final recommended MSD value for each vignette.

**Results:** Final MSD ratings for the five subscales of Bayley-4 and 10 subscales of the Vineland-3 had an agreement rate of 70% or higher. MSD thresholds for each subscale were not single cut-offs, but rather reflected a range of MSD values dependent on level of baseline functioning.

**Conclusions:** The Delphi Panel method incorporates the caregiver perspective to provide preliminary estimates of what constitutes meaningful within person change on the Bayley-4 and Vineland-3 in individuals with AS with various levels of baseline functioning.

**Highlights:**

⍰ To acquire regulatory approval in drug development, sponsors must demonstrate both statistical significance and clinical meaningfulness of a treatment effect.While several clinical trials are underway in AS, within person meaningful score difference thresholds are not yet established for the most commonly used outcome measures, namely the Bayley and Vineland.
⍰ Aligning with FDA guidance, we have developed an innovative qualitative approach using a Delphi panel to incorporate caregiver perspectives in defining meaningful change and generated preliminary patient-informed meaningful score differences (MSDs) for individuals with Angelman Syndrome.
⍰ What caregivers of individuals with AS consider to be a MSD on the measures depends primarily on the baseline severity of their child’s presentation.

## Introduction

Angelman syndrome (AS) is a rare neurodevelopmental disorder with an estimated prevalence of approximately 1:22,000 to 1:52,000.^1–3^ Individuals with AS have severe global developmental delay leading to intellectual disability, minimal expressive language, ataxia, seizures, challenging behaviors, and sleep disturbances. Adults with AS require supportive living and full-time assistance with all activities of daily living.^4, 5^ AS is caused by a lack of expression of the maternally-inherited copy of *UBE3A* in neurons, which may be due to a deletion on chromosome 15, i.e., “deletion positive” or other genetic etiologies not due to a deletion, i.e., “deletion negative.”^6–8^ Deletion-positive individuals are more developmentally delayed than deletion-negative individuals.^9–12^

Currently, there is no disease-modifying treatment that targets the underlying pathophysiology of AS.^13, 14^ However, several pharmaceutical companies are testing novel therapeutic strategies to directly restore or replace *UBE3A* in the brain using antisense oligonucleotides^15–18^ or gene therapy. In these clinical trials, measures assessing development (e.g., Bayley Scales of Infant and Toddler Development^19^) and adaptive functioning (e.g., Vineland Adaptive Behavior Scales^20^) are among the primary and secondary outcome measures. To achieve regulatory approval, both the statistical and clinical significance of the effect of intervention must be demonstrated.^21^ While clinical meaningfulness has been examined in other neurodevelopmental disorders^22–26^, none are specific to individuals with AS. Thus, the minimum change in the scores that should be considered “clinically meaningful” in AS for common outcome measures is unknown, making it difficult to define “success” and assess how clinically meaningful the observed treatment effect is in clinical trials.^27^

One approach to support clinical significance is to define “meaningful score difference” (MSD), i.e., the minimum within-person change score on a clinical outcome assessment (COA) that would be perceived as clinically meaningful for patients or their caregivers.^28^ Although there are multiple approaches to defining MSDs in a clinical trial, the FDA encourages the use of external anchors in which changes in the COA scores are “anchored” against one or more external criteria using established measures of clinical relevance, such as a global impressions rating of symptom severity.^23^ However, in the absence of an appropriate external measure collected in conjunction with the COAs to anchor MSDs, an alternative approach is needed. In this study, we utilized a novel, stakeholder-informed, qualitative approach to the development of MSDs. Specifically, we asked caregivers and clinicians to provide input, through a formal Delphi panel approach, on what would constitute a meaningful change on the Bayley and the Vineland scales. We then quantified their responses in order to establish preliminary MSDs, which sponsors of future clinical trials in AS can use as a foundation for establishing clinical meaningfulness.

## Methods

A Delphi panel comprised of caregivers was used to gather data on MSDs for each subscale of the Bayley-4 and Vineland-3. Since individuals with AS have severe cognitive and verbal limitations, caregivers served as surrogate informants. MSDs may be dependent on multiple patient factors including age, and molecular subtype (deletion positive and deletion negative), which could make a single MSD for a given outcome measure inappropriate. ^9–12,28^ We therefore recruited caregivers to represent these criteria. This study was approved by the institutional review board at [BLINDED FOR REVIEW].

### Participants

The Delphi panel consisted of a convenience sample of 19 caregivers of individuals who participated in the AS Natural History Study (ASNHS)^29^, in the following age and subtype categories: deletion-positive younger than 5 years (DP<5yrs), *n* = 4; deletion-positive 5 years or older (DP≥5yrs), *n* = 5; deletion-negative younger than 5 years (DN<5yrs), *n* = 4; and deletion-negative 5 years or older (DN≥5yrs), *n* = 6.

Five highly-experienced clinicians were also included in the Delphi panel meetings. Caregivers are considered experts in the lived experience of AS and the closest proxy to patients’ perspectives; therefore, the final MSD results reflect only these perspectives. Including both caregivers and clinicians in the discussion of meaningful change provides a more holistic approach—with caregivers providing highly personalized meaningful change ratings based on their lived experiences, complemented by clinicians who have a broader experience with a large number of patients across age and deletion subtypes. Consistent MSD ratings between caregivers and clinicians would further support the robustness of the primary results obtained. Caregiver and clinician characteristics are summarized in Table 1.

**Table 1.** Characteristics of the Delphi Panel.

| **Caregiver Characteristics** |  |
| --- | --- |
| Survey Respondents | 17 |
| Race/Ethnicity |  |
| Black/African American | 1 (6%) |
| Hispanic/Latino | 2 (12%) |
| Non-Hispanic White | 14 (82%) |
| Educational Attainment |  |
| High School or Less | 1 (6%) |
| Some College | 6 (33%) |
| College Degree | 4 (22%) |
| Graduate Degree or Higher | 6 (33%) |
| US Regional Representation |  |
| Midwest | 4 (24%) |
| Northeast | 5 (29%) |
| South | 7 (41%) |
| West | 1 (6%) |
| Number of Children with AS |  |
| One Child | 16 (94%) |
| Two Children | 1 (6%) |
| Caregiver Status |  |
| Father | 3 (16%) |
| Mother* | 16 (84%) |
| Primary Caregiver | 14 (82%) |
| AS Subtype of Children |  |
| Deletion-Positive | 9 (53%) |
| Deletion-Negative | 8 (47%) |
| Age of Children in Years |  |
| At Diagnosis |  |
| Median (IQR) | 1.7 (0.75) |
| Range | (0.75-10) |
| At Start of Panel Meetings |  |
| Median (IQR) | 5 (14) |
| Range | (2-35) |
| Instrument Familiarity |  |
| Observed Bayley administration | 12 (71%) |
| Participated in Vineland administration | 12 (71%) |
| Experience with either a Bayley or a Vineland administration | 14 (82%) |
| Experience with administrations of both Bayley and Vineland | 10 (59%) |
| **Clinician Characteristics** |  |
| Survey Respondents | 5 |
| Race/Ethnicity |  |
| Asian | 1 (20%) |
| Non-Hispanic White | 4 (80%) |
| Clinician Specialization |  |
| Clinical Geneticist | 3 (60%) |
| Neurologist | 1 (20%) |
| Psychologist | 1 (20%) |
| AS Experience |  |
| Years working with AS patients |  |
| Median (IQR) | 22 (13) |
| Range | (10-35) |
| Number of AS patients seen annually |  |
| Median (IQR) | 30 (45) |
| Range | (28-175) |
| Clinical Setting |  |
| Academic medical center | 5 (100%) |
| Instrument Familiarity |  |
| Administered Bayley | 0 (0%) |
| Administered Vineland | 2 (40%) |
| Observed either a Bayley or a Vineland administration | 5 (100%) |
| Observed both Bayley and Vineland administrations | 3 (60%) |

### Instruments, Subscales, and Metrics

The Bayley-4^19^ is a clinician led, direct assessment of early developmental skills, normed for children aged 16 days to 42 months. It is frequently used out of age range for individuals with AS.^9, 12^ The Bayley-4 assesses skills in five subscales: Gross Motor, Fine Motor, Receptive Communication, Expressive Communication, and Cognition; each contains items measuring specific skills with an item score of ‘0’ indicating no achievement, ‘1’ indicating partial achievement, and ‘2’ indicating full achievement of the skill.

The Vineland-3^20^ is a widely used assessment of adaptive functioning, i.e., how well an individual regularly uses specific skills in their daily life to achieve age-appropriate levels of independence. The Comprehensive Interview version, which is a semi-structured caregiver interview conducted by experienced clinicians, was used in this study. The Vineland-3 comprises eleven subscales that are organized within four domains: Communication (Expressive, Receptive, Written), Daily Living (Personal, Domestic, Community), Socialization (Interpersonal Relationships, Playtime and Leisure, Coping), and Motor (Gross and Fine). In this study, all subscales were included except ‘Written’, which reflects early academic skills that are not typically attained by individuals with AS. For each subscale, the Vineland-3 assesses skills of increasing complexity where an item score of 0 indicates that the individual does not perform a skill independently, 1 indicates they sometimes perform it independently, and 2 indicates they usually perform it independently.

#### Scoring metrics

Bayley-4 and Vineland-3 are standardized norm-referenced assessments and offer a range of scoring metrics, including growth scale values (GSVs). GSVs are a Rasch model-based transformation of the raw score that have an interval measurement level and a conditional standard error of measurement (CSEM)^30^ and are therefore intended for monitoring change.^17,18^ Previous studies have shown that GSVs are reasonably sensitive to capture change in individuals with AS.^11, 12^ GSVs are available, but standard scores are not, when an instrument is used out of the age range of the norm sample. Therefore, we focused on establishing MSDs based on GSVs for both the Bayley-4 and Vineland-3.

### Vignettes

Two to four vignettes of varying baseline functional levels were developed for each of the five Bayley-4 and 10 Vineland-3 subscales (N=43 vignettes) (see Figure 1 for example). Each vignette presented a hypothetical individual with AS, describing their baseline set of skills or behaviors on the subscale and showing the next most difficult skills or behaviors in order of increasing difficulty. The skills and behaviors reflected the content of items from the Bayley-4 and Vineland-3. For the purpose of this exercise, it was assumed that each individual would have full scores on all items up to and including a particular item, and scores of zero on the remaining items (consistent with the instruments’ basal and ceiling rules). The raw score associated with each such pattern of item scores was converted to a GSV. This enabled estimation of the GSV increase associated with the acquisition of additional, more difficult skills or behaviors. Each such increase was a candidate MSD.

**Figure 1.**
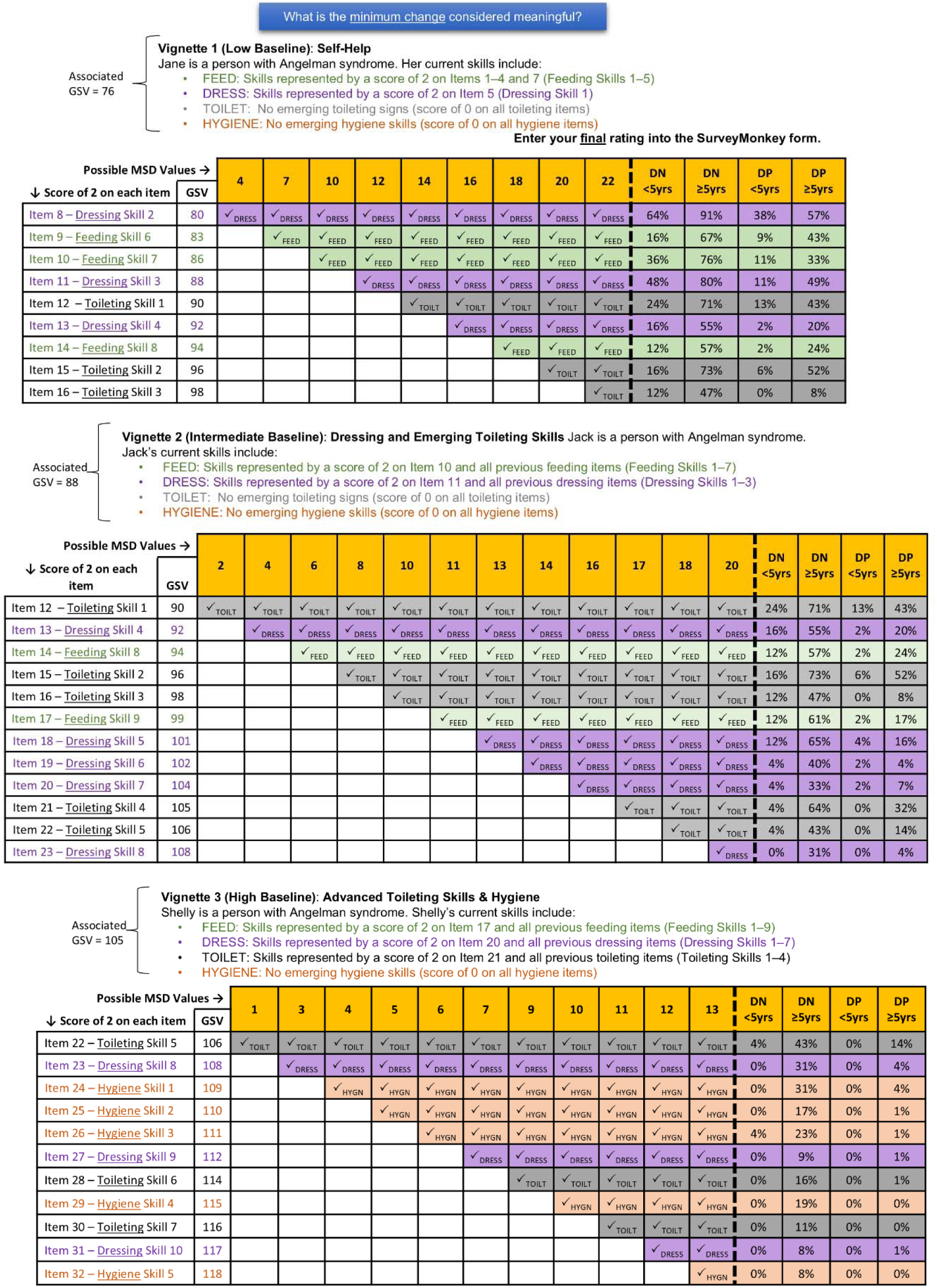
Vineland Personal Subscale Vignettes Figure 1 footnote: The low baseline vignette includes a baseline skill level described by full credit on items 1-7. The associated GSV for this baseline is 76. Items 8-16 are included as possible MSD values above baseline. Consistent achievement (a score of 2) of the skills measured by items 1-8, for example, results in a GSV score of 80. The change from baseline would therefore be 80-76, i.e., 4 (the value indicated in column 1). If an MSD of 10 was selected, this would mean that an individual at a baseline GSV of 76 would demonstrate a meaningful score difference if they were later able to consistently achieve the skills in items 8 (an item measuring the second dressing skill in the assessment), 9 (an item measuring the sixth feeding skill on the assessment), and 10 (an item measuring the seventh feeding skill on the assessment).

Baseline functional levels for each vignette were set to be in the range of GSVs currently obtainable by individuals with AS, as determined from extant GSV data from the ASNHS^21, 29^ and a longitudinal study focused on endpoint development (FREESIAS)^31^ (Bayley-III: *n* = 531; Vineland-3: *n* = 526). Limited Bayley-4 data were available at the time of the study so item level data from the Bayley-III was used and cross walked to corresponding items of the Bayley-4. For most subscales, vignettes were created for a low, medium, and high baseline functional level. For subscales in which few items separated the GSV scores for the “low” and “high” baselines, the “medium” category was omitted. The percentage of individuals with AS who achieved each candidate MSD was also calculated using these data.

A wide range of potential MSDs was included for each vignette to ensure that the panelists’ ratings were not constrained to a range of score differences that might be too small to be considered meaningful. MSDs did not exceed the growth a typically developing child would be expected to make in one year based on the instrument-supplied age equivalents—as a change that large for individuals with AS should be considered meaningful.

### Delphi Panel Design

#### Instructions to Panelists

Panelists were asked to identify the smallest change that they would consider to be meaningful based on the vignette they were presented with. We developed the following definition of meaningful change, taking into account FDA guidance^28^:

> *The degree or level of change in a measure that the caregiver feels noticeably impacts their child’s daily functioning or family quality of life in a way that is important to the child or family*.

This definition was reinforced throughout the study to the panelists. All panelists were instructed to consider meaningful change based on their own experiences and the descriptions provided in the vignettes. They were specifically asked to consider if meaningful change varied based on factors such as baseline skill level, age, or AS subtype. The panelists were told not to consider the rate at which the change would occur or the context of any treatments and associated risks/side effects. Caregivers were asked to evaluate the vignettes for their particular group (DP<5yrs, DP≥5yrs, DN<5yrs, DN≥5yrs). Clinicians were asked to consider what they thought families would notice and find meaningful when making judgments about MSDs and asked to provide ratings for all four AS groups.

#### Delphi Panel Meeting Structure

Ten virtual Delphi panel meetings were held over the course of six months. The first meeting provided training and piloted the process using the Vineland-3 Personal subscale. The remaining meetings included collection of three rounds of independent panelist ratings for each of the 15 subscales.

For Round 1, panelists received the vignettes and completed ratings independently in advance of the panel meeting using SurveyMonkey (https://www.surveymonkey.com/) so the ratings could be compiled and caregiver agreement calculated. Vignettes were presented in order from the lowest to highest baseline skill attainment. During the Delphi panel meetings, panelists were assigned to one of three breakout rooms facilitated by a member of the research team: one breakout room for the clinicians; another for the DN caregivers (DN<5yrs and DN≥5yrs); and a third for the DP caregivers (DP<5yrs and DP≥5yrs). This prevented caregivers from being overly influenced by the opinions of clinicians and provided an opportunity to obtain potentially different definitions of meaningful change among the DP and DN groups. Each facilitator in a breakout room led a discussion of Round 1 ratings during which the panelists in the room described their rationale for their ratings, listened to the rationales of other panelists, and discussed differences in ratings with the goal of moving toward consensus.

After the panelists completed discussion of their Round 1 ratings, each panelist independently submitted their Round 2 ratings. After the meeting, the agreement rate among caregivers was updated based on Round 2 results. In a subsequent meeting, a facilitator asked panel members to reflect upon the aggregated Round 2 ratings.

When Round 2 ratings across all panelists were consistent, Round 3 was used to collect a final yes/no endorsement of the median group rating from each panelist. If a panelist did not endorse the median group rating, they were asked to provide a rationale. If Round 2 ratings were still inconsistent, individual Round 3 ratings were collected using the same process used in previous rounds. Round 3 was the final round regardless of whether 100% agreement was reached. The iterative Delphi panel process was intended to build consensus. However, because people may disagree about the amount of change that they consider meaningful, consensus was not required. Panelists were also surveyed periodically about their satisfaction with the Delphi process. The full list of questions posed to the panel is provided in the Supplemental materials.

### Calculating Meaningful Score Differences

Following formal Delphi panel procedures^32^, the median rating from caregivers was retained as the final recommended MSD value for each vignette. Technically, each MSD is associated with a single baseline GSV. However, we provided an initial recommendation for an expanded range of baseline GSVs for which an MSD might be applicable so that the MSDs might be considered with a wider range of baseline functional levels. The expanded GSV range was created using the midpoints between baseline GSVs and the region of the scale with conditional standard errors of measurement (CSEM) no larger than 0.5 more than the CSEM at the baseline GSV. This CSEM threshold was used as a guardrail because we have less confidence about the applicability of MSD values recommended by the Delphi panel at the extreme ends of the scale where measurement error is substantially higher.

### Comparative Metrics

To contextualize the magnitude of caregiver MSDs, they were compared to several other change metrics^33^: 0.5 standard deviations of the baseline GSV distribution for each AS group, expected GSV change in 12 months for typically developing individuals (based on age equivalents from the publisher) and individuals with AS (based on NHS and FREESIAS data), and the reliable change index (RCI) for GSVs. Details on the calculation of these metrics are provided in the Supplement.

### Ratings of Subscale and Skill Relevance

We asked caregivers and clinicians to rate the importance of each subscale for inclusion as outcome measures in future AS clinical trials. Additionally, because we observed that some skills within a subscale were considered more important to caregivers during Delphi panel discussions, we asked panelists to rate the importance of skills within each subscale.

## Results

### Meaningful Score Differences

Given our hypothesis that MSDs differ by age and molecular subtypes (DN<5yrs, DN≥5yrs, DP<5yrs, DP≥5yrs), we compared results by group before determining whether ratings could be combined (see Tables 2A-2C). Contrary to our hypothesis, a comparison of median MSDs generally indicated no differences among the groups, and where differences were found, the patterns were not consistent. Because age and subtypes had little to no influence on panelists’ ratings, the final median MSDs were calculated from the ratings of all four caregiver groups. However, different MSD values were obtained for different baseline functional levels, suggesting that meaningful change is dependent upon baseline function.

**Table 2A.**
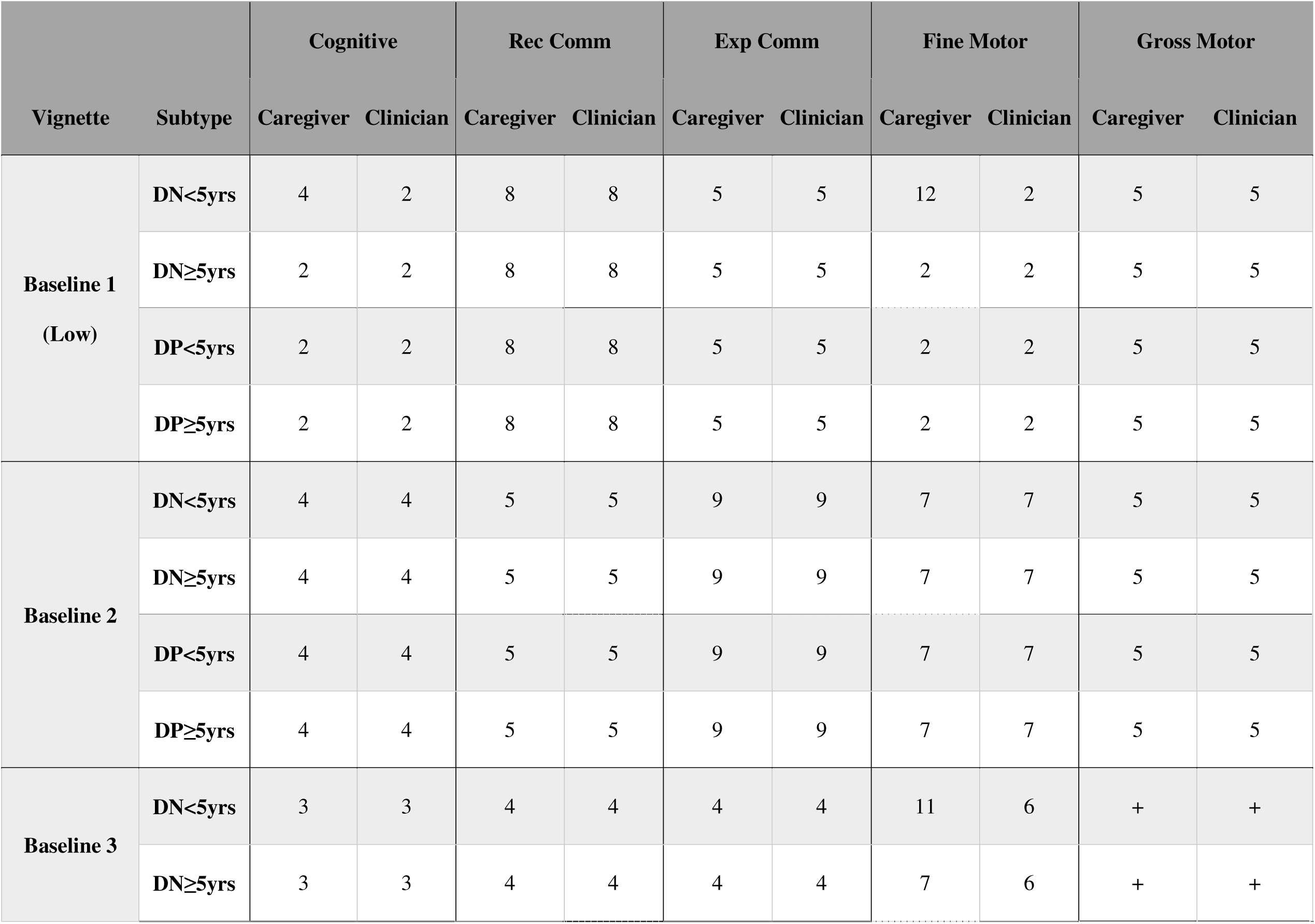
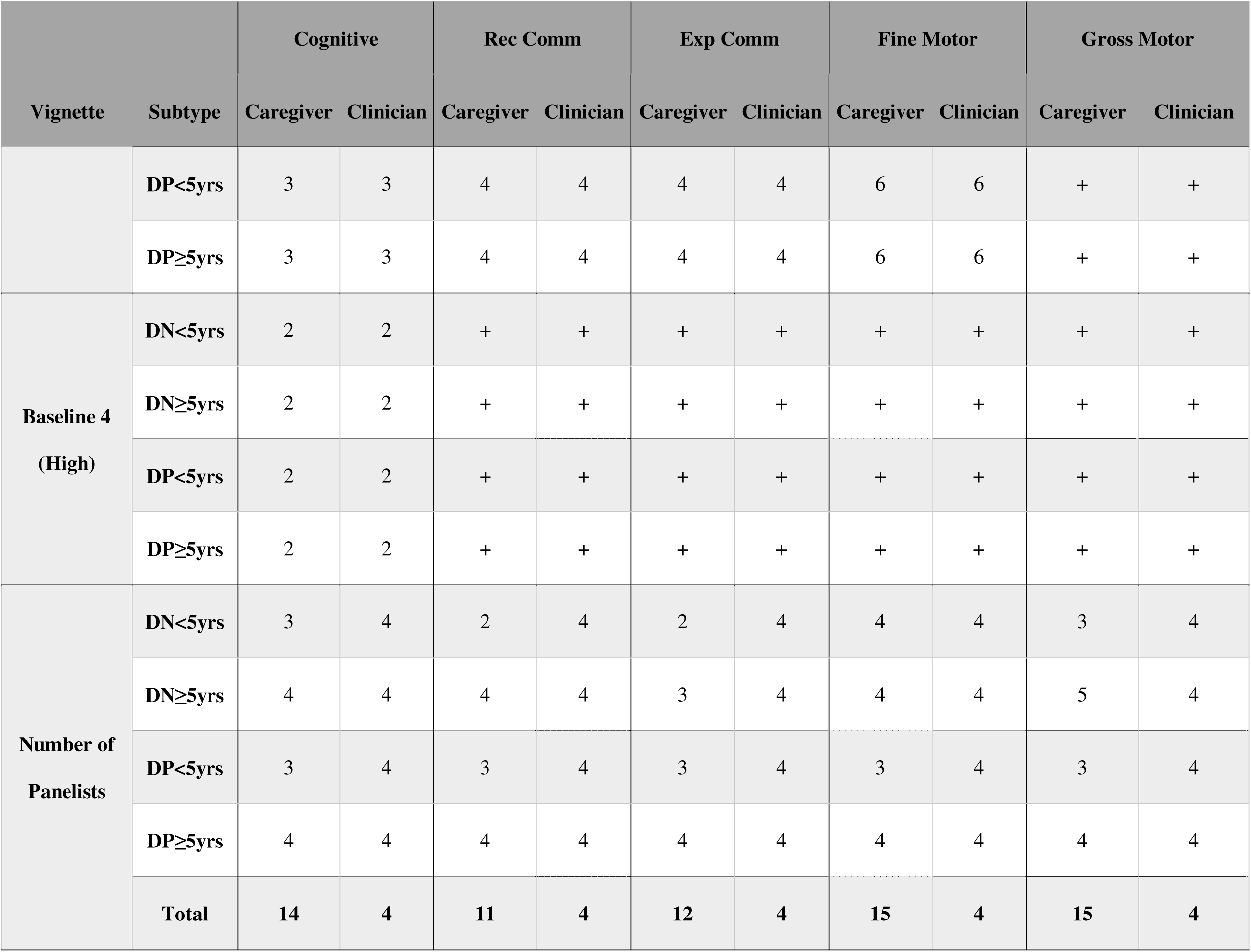
Median GSV Meaningful Score Differences by Molecular Subtype and Age (Bayley-4)

**Table 2B.**
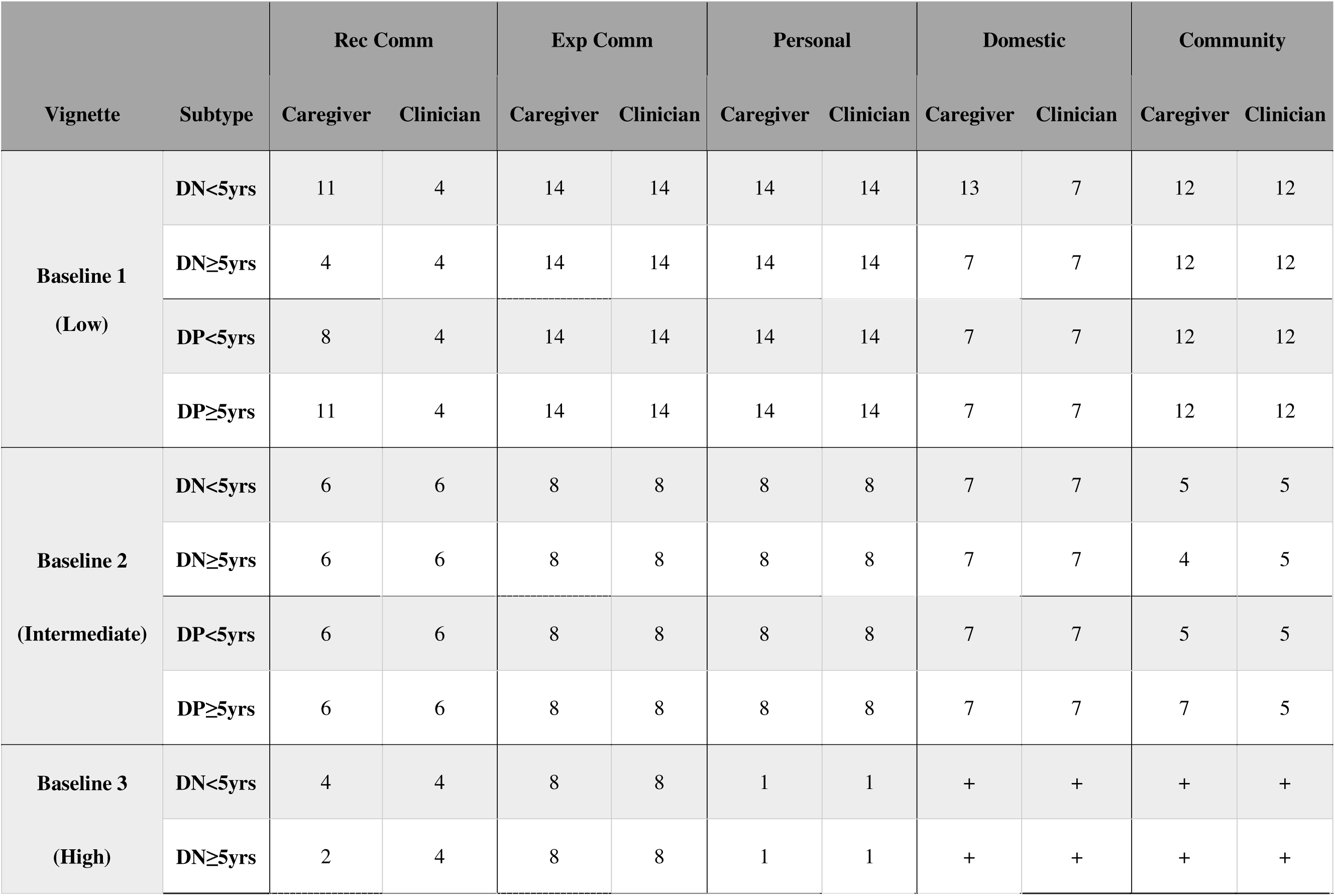
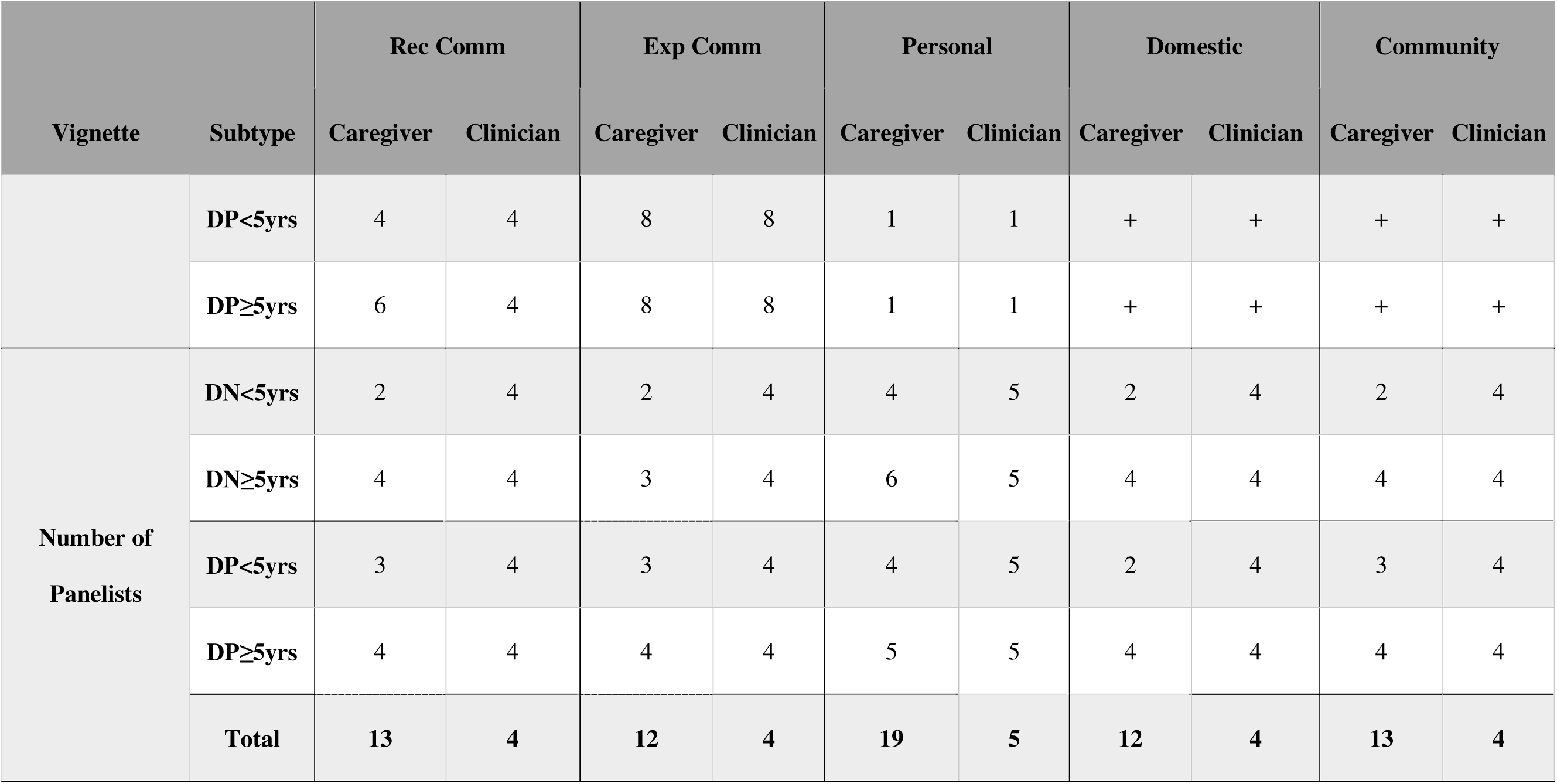
Median GSV Meaningful Score Differences by Molecular Subtype and Age (Vineland-3)

**Table 2C.** Median GSV Meaningful Score Differences by Molecular Subtype and Age (Vineland-3)

|  |  | Interpersonal |  | Play & Leisure |  | Coping |  | Gross Motor |  | Fine Motor |  |
| --- | --- | --- | --- | --- | --- | --- | --- | --- | --- | --- | --- |
| Vignette | Subtype | Caregiver | Clinician | Caregiver | Clinician | Caregiver | Clinician | Caregiver | Clinician | Caregiver | Clinician |
| Baseline 1<br>(Low) | DN<5yrs | 7 | 7 | 22 | 22 | 5 | 4 | 24 | 24 | 20 | 20 |
|  | DN≥5yrs | 7 | 7 | 22 | 22 | 4 | 4 | 24 | 24 | 20 | 20 |
|  | DP<5yrs | 7 | 7 | 20 | 22 | 4 | 4 | 24 | 24 | 18 | 20 |
|  | DP≥5yrs | 7 | 7 | 22 | 22 | 6 | 4 | 24 | 24 | 20 | 20 |
| Baseline 2<br>(Intermediate) | DN<5yrs | 4 | 4 | 2 | 3 | 4 | 2 | 10 | 7 | 10 | 10 |
|  | DN≥5yrs | 4 | 4 | 2 | 3 | 4 | 2 | 11 | 7 | 10 | 10 |
|  | DP<5yrs | 4 | 4 | 2 | 3 | 3 | 2 | 7 | 7 | 10 | 10 |
|  | DP≥5yrs | 4 | 4 | 4 | 3 | 8 | 2 | 14 | 7 | 10 | 10 |
| Baseline 3 | DN<5yrs | 1 | 1 | 3 | 3 | 4 | 3 | 7 | 7 | 9 | 11 |

Meaningful Change in Angelman Syndrome
|  |  | Interpersonal |  | Play & Leisure |  | Coping |  | Gross Motor |  | Fine Motor |  |
| --- | --- | --- | --- | --- | --- | --- | --- | --- | --- | --- | --- |
| Vignette | Subtype | Caregiver | Clinician | Caregiver | Clinician | Caregiver | Clinician | Caregiver | Clinician | Caregiver | Clinician |
| (High) | DN≥5yrs | 1 | 1 | 3 | 3 | 3 | 3 | 7 | 7 | 11 | 11 |
|  | DP<5yrs | 1 | 1 | 3 | 3 | 1 | 3 | 7 | 7 | 8 | 11 |
| | DP≥5yrs | 1 | 1 | 3 | 3 | 3 | 3 | 7 | 7 | 11 | 11 |
| Number of Panelists | DN<5yrs | 2 | 4 | 2 | 4 | 3 | 4 | 3 | 4 | 2 | 4 |
| | DN≥5yrs | 3 | 4 | 3 | 4 | 4 | 4 | 5 | 4 | 4 | 4 |
|  | DP<5yrs | 2 | 4 | 2 | 4 | 3 | 4 | 3 | 4 | 3 | 4 |
| | DP≥5yrs | 3 | 4 | 3 | 4 | 4 | 4 | 4 | 4 | 4 | 4 |
|  | Total | 10 | 4 | 10 | 4 | 14 | 4 | 15 | 4 | 13 | 4 |

The Delphi panel results are summarized in terms of final median caregiver MSD ratings and rater agreement in Table 3. As an illustration of how to interpret the values in Table 3, consider the low baseline for the Bayley-4 Gross Motor subscale. This vignette was developed using a baseline GSV of 491 and may be applicable to extended baseline GSVs of 462–501 as this range reflects lower gross motor function and has similar measurement error. After three rounds of ratings, caregivers unanimously rated five GSV points as the MSD. Thus, individuals with baseline GSVs between 462 and 501 would need to gain skills associated with at least five GSV points to demonstrate a MSD.

**Table 3.** Meaningful Score Differences by Vignette.

| **Subscale** | **Vignette**<br>**(Baseline**<br>**Performance)** | **Extended**<br>**Baseline**<br>**GSV Range<sup>1</sup>** | **MSD Rating**<br>**[MSD Range]<sup>2</sup>** | **Percent**<br>**Agreement after**<br>**Round 3** |
| --- | --- | --- | --- | --- |
| **Bayley-4<sup>3</sup>** |  |  |  |  |
| Cognitive | Low (GSV = 492) | 459-497 | 2 | 73% |
|  | Int Low (GSV = 503) | 498-508 | 4 | 100% |
|  | Int High (GSV = 514) | 509-519 | **3** [2-3] | 91%<sup>4</sup> |
|  | High (GSV = 525) | 520-557 | 2 | 75% |
| Rec Comm | Low (GSV = 497) | 451-501 | 8 | 100% |
|  | Int (GSV = 505) | 502-512 | 5 | 100% |
|  | High (GSV = 520) | 513-556 | 4 | 100% |
| Exp Comm | Low (GSV = 485) | 477-491 | 5 | 100% |
|  | Int (GSV = 498) | 492-505 | 9 | 100% |
|  | High (GSV = 510) | 506-545 | 4 | 100% |
| Fine Motor | Low (GSV = 491) | 455-497 | 2 | 73% |
|  | Int (GSV = 503) | 498-511 | 7 | 100% |
|  | High (GSV = 520) | 512-549 | **6** [6-8] | 82%<sup>4</sup> |
| Gross Motor | Low (GSV = 491) | 462-501 | 5 | 100% |
|  | High (GSV = 512) | 502-539 | 5 | 100% |
| **Vineland-3** |  |  |  |  |
| Rec Comm | Low (GSV = 55) | 52-66 | **11** [4-11] | 100%<sup>4</sup> |
|  | Int (GSV = 79) | 67-91 | 6 | 80% |
|  | High (GSV = 104) | 92-127 | **4** [2-6] | 90%<sup>4</sup> |
| Exp Comm | Low (GSV = 65) | 61-82 | 14 | 100% |
|  | Int (GSV = 99) | 83-105 | 8 | 100% |
|  | High (GSV = 112) | 106-167 | 8 | 100% |
| Personal | Low (GSV = 76) | 70-81 | 14 | 100% |
|  | Int (GSV = 88) | 82-96 | 8 | 100% |
|  | High (GSV = 105) | 97-136 | 1 | 79% |
| Domestic | Low (GSV = 27) | 27-31 | 7 | 80% |
|  | High (GSV = 38) | 32-85 | 7 | 90% |
| Community | Low (GSV = 25) | 25-31 | 12 | 100% |
|  | High (GSV = 39) | 32-109 | **5** [5-7] | 78%<sup>4</sup> |
| Interpersonal | Low (GSV = 58) | 42-71 | 7 | 100% |
|  | Int (GSV = 85) | 72-92 | 4 | 90% |
|  | High (GSV = 99) | 93-118 | 1 | 100% |
| Play &<br>Leisure | Low (GSV = 54) | 54-68 | 22 | 80% |
|  | Int (GSV = 83) | 69-89 | **2** [2-4] | 100%<sup>4</sup> |
|  | High (GSV = 95) | 90-112 | 3 | 100% |
| Coping | Low (GSV = 45) | 41-51 | **4** [4-6] | 91%<sup>4</sup> |
|  | Int (GSV = 58) | 52-62 | **5** [2-8] | 100%<sup>4</sup> |
|  | High (GSV = 66) | 63-82 | **3** [1-3] | 82%<sup>4</sup> |
| Gross Motor | Low (GSV = 62) | 50-81 | 24 | 91% |
|  | Int (GSV = 103) | 82-114 | **7** [7-14] | 73%<sup>4</sup> |
|  | High (GSV = 125) | 115-149 | 7 | 100% |
| Fine Motor | Low (GSV = 78) | 57-92 | 20 | 90% |
|  | Int (GSV = 105) | 93-110 | 10 | 100% |
|  | High (GSV = 118) | 111-157 | **11** [6-11] | 100%<sup>4</sup> |

Based on data in Table 3, agreement on a single MSD value was 70% or higher for 31 of 43 (72%) of vignettes, including 100% agreement on 21 of 43 (49%). In cases where caregivers’ final ratings did not reach 70% agreement (12 of 43 vignettes, 28%), a MSD range was provided in addition to the median MSD value. Any rating endorsed by at least two caregivers was included in the MSD range. For example, the High/Intermediate baseline for the Cognitive subscale had two modal MSD ratings (2 and 3), each endorsed by 5 of 11 caregivers. Thus, a range of 2-3 was defined as the MSD (Table 3), with the median value of 3 emphasized in bold font to reflect a more conservative criterion: AS individuals making this amount of change from a baseline GSV of 514 will meet or exceed the threshold considered meaningful by 91% of the caregivers included in this study’s Delphi panel.

A comparison of caregiver MSD ratings with clinician ratings indicated highly consistent results. In fact, clinician and caregiver MSDs matched in 42 out of 43 vignettes. The single exception was the low/intermediate baseline for the Bayley-4 Cognitive subscale where the caregiver median rating was four and the clinician median rating was two.

### Comparative Metrics

A comparison of MSD values to other metrics of change (see Vineland-3 and Bayley-4 Distribution-Based Summary Tables in the Supplement) indicated that MSD values often exceeded the typical change expected over 12 months in an individual with AS, especially for the deletion and non-deletion groups with age ≥ 5 years. MSDs were always less than the 12-month change for a typically developing individual. The MSDs were similar to the 0.5SD and RCI values.

### Subscale and Skill Importance

For clinicians and all four groups of caregivers, Communication, Gross Motor, Cognitive, and Interpersonal Relationships and Coping Socializations skills were rated of high importance for inclusion as outcomes in clinical trials. Domestic, Play and Leisure, and Fine Motor skills were rated among the least important. Clinicians rated Cognitive and Personal Daily Living skills more highly than caregivers (see Figure 2).

**Figure 2.**
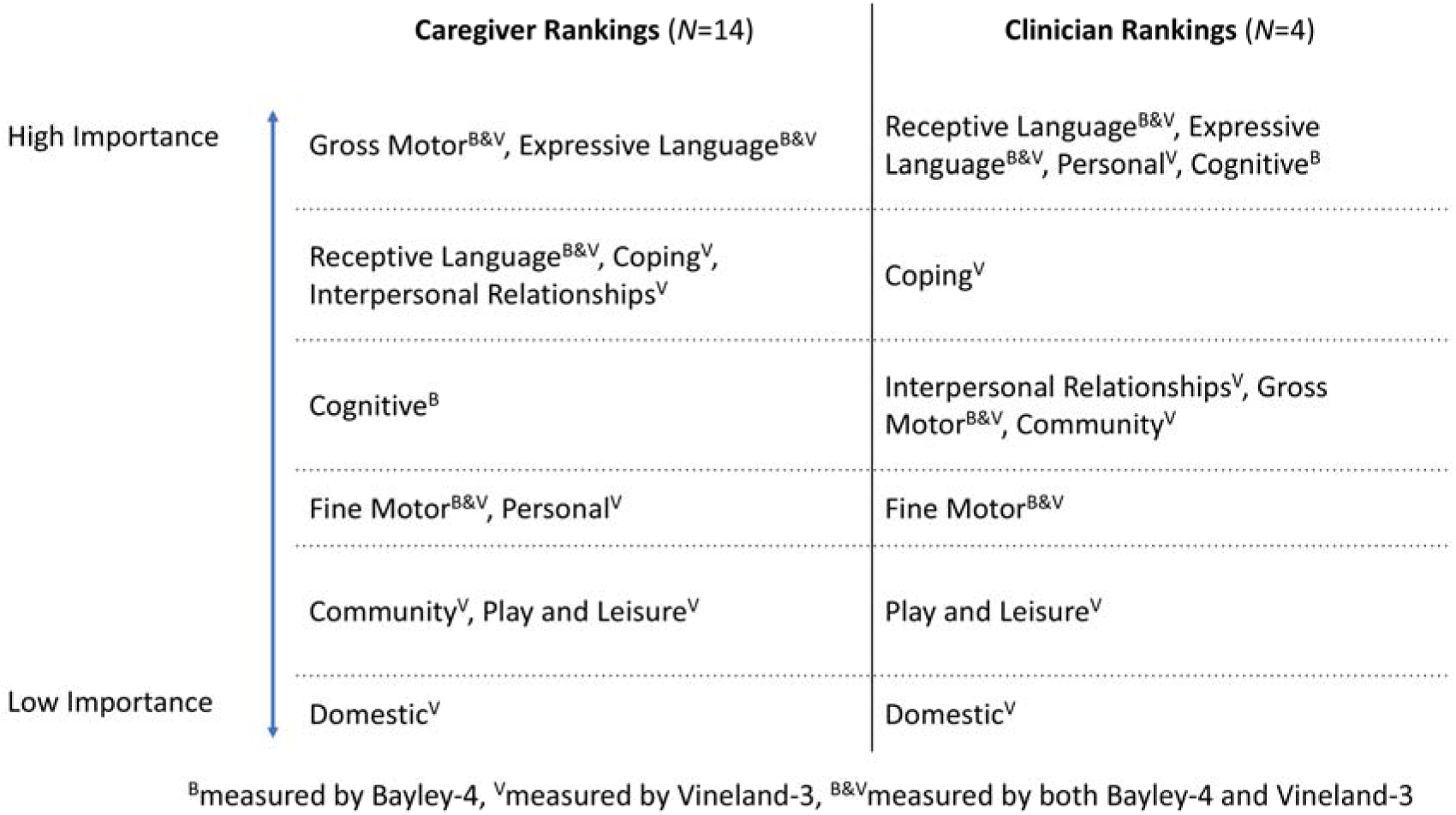
Subscale Importance as Rated by Caregivers and Clinicians

Caregivers and clinicians often agreed on the relative importance of a skill within a subscale. For example, within the Personal subscale, feeding and toileting were rated as more important while dressing and hygiene were rated as less important by both caregivers and clinicians. Within the Gross Motor subscale, clinicians rated skills related to balance as being more important than caregivers did, although in discussions, caregivers also noted the importance of skills related to balance for safety. Skill importance ratings for caregivers and clinicians are provided in Tables 4 and 5. Panelists frequently mentioned skills that increased the safety or prevented injury of their child as being very important to quality of life; self-entertainment and other skills that enabled caregivers to reduce their vigilance and attend to other household needs were also a focus of discussions of meaningful change. Panelists also noted that what is most important to one family may not be what is most important to another family, which complicates having a single threshold to use for evaluation of therapeutic interventions (see Table 5 in the Supplement). Survey results (provided in detail in the Supplementary materials) indicated that caregivers understood the process and felt confident in the final results.

**Table 4.**
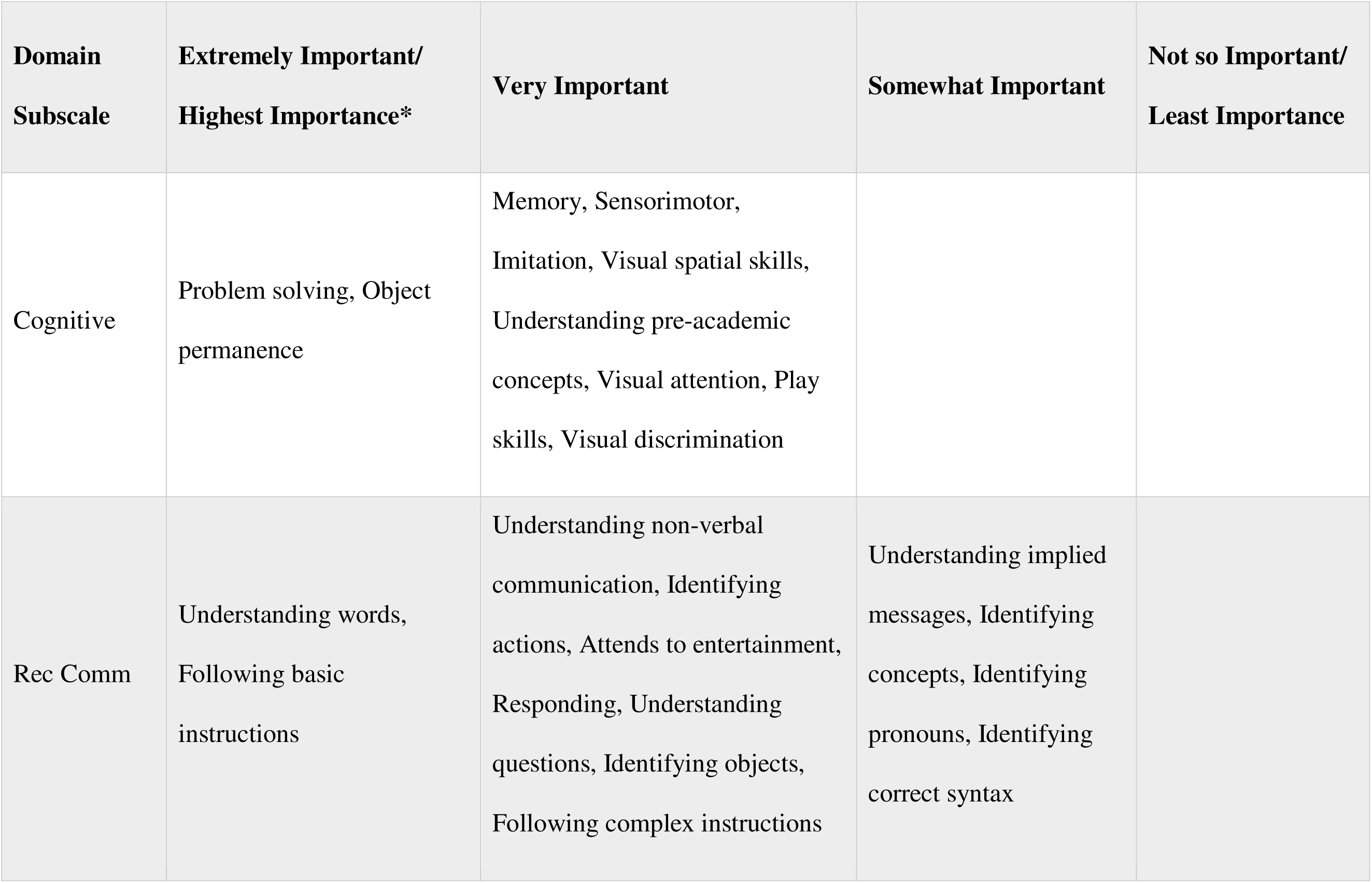
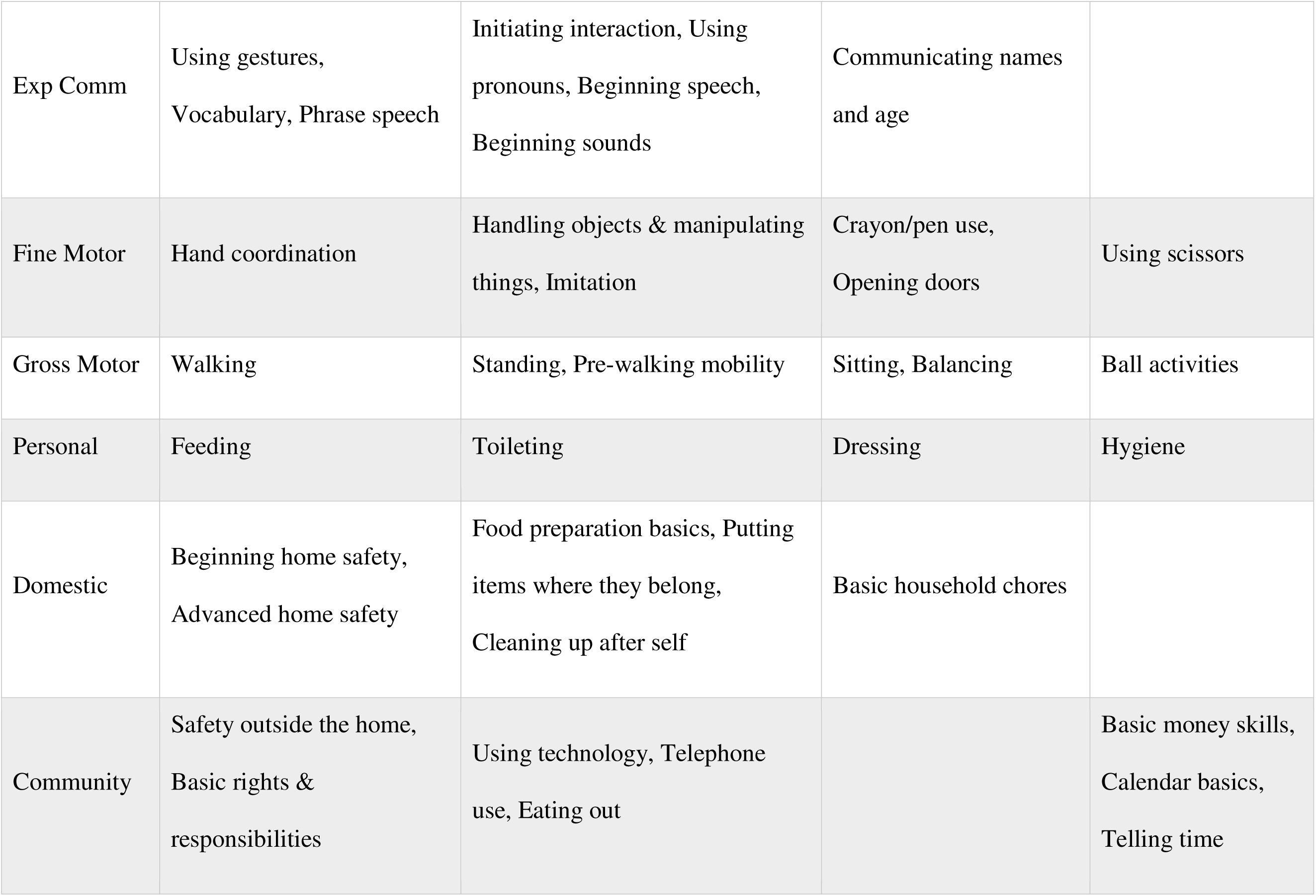
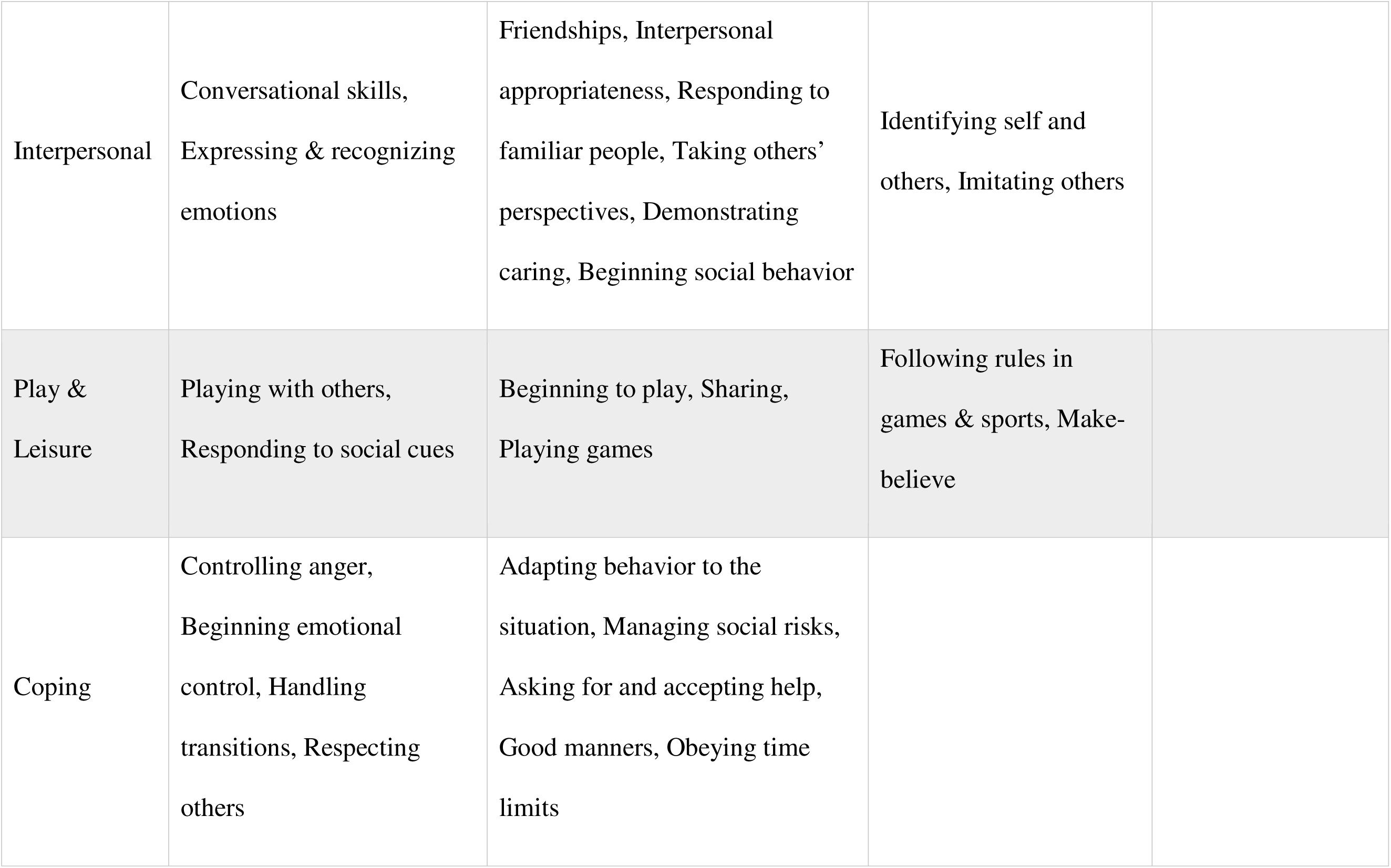
Ratings of the Relevance of Within-Subscale Skills (Caregiver Results)

**Table 5.**
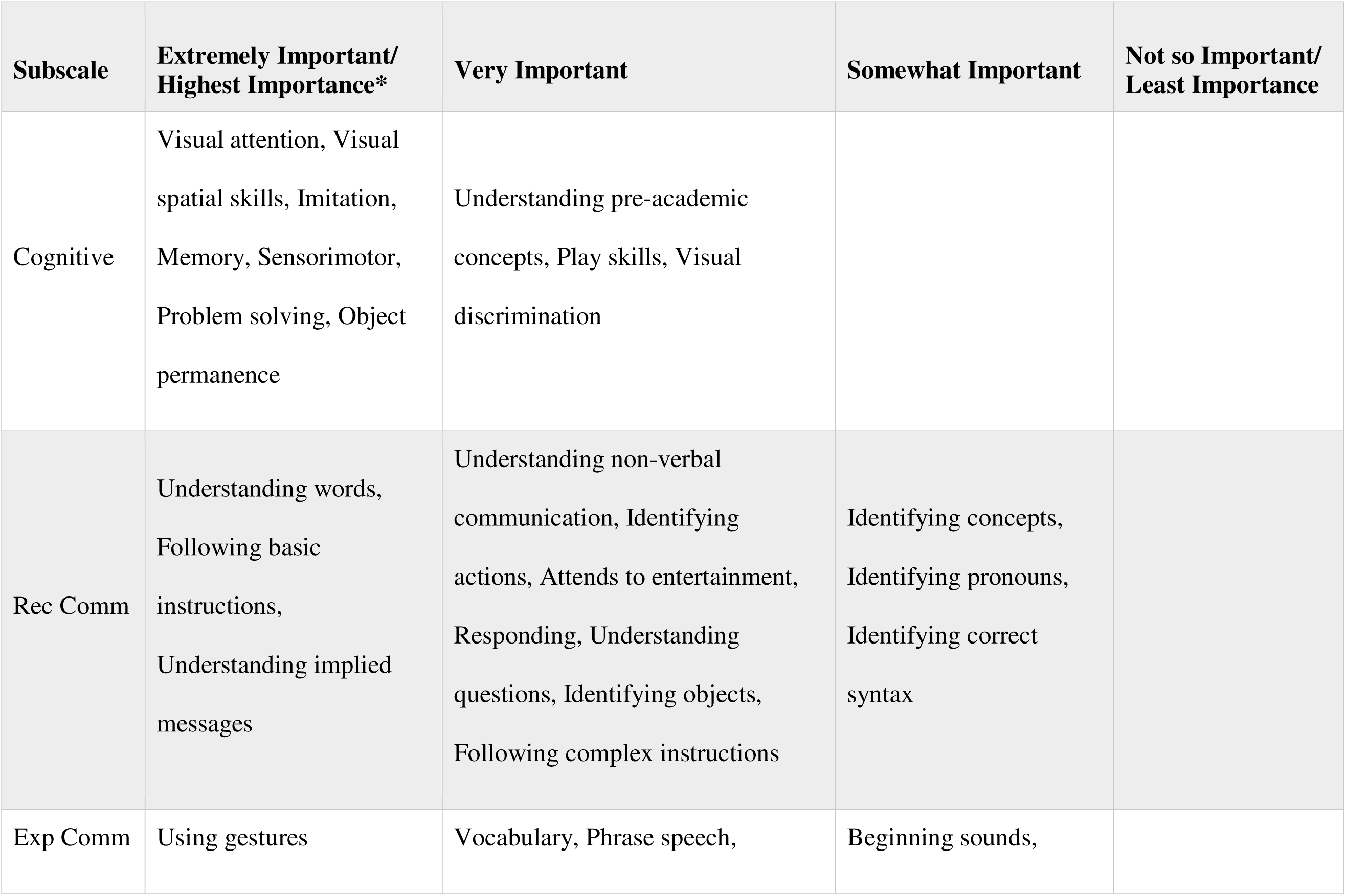
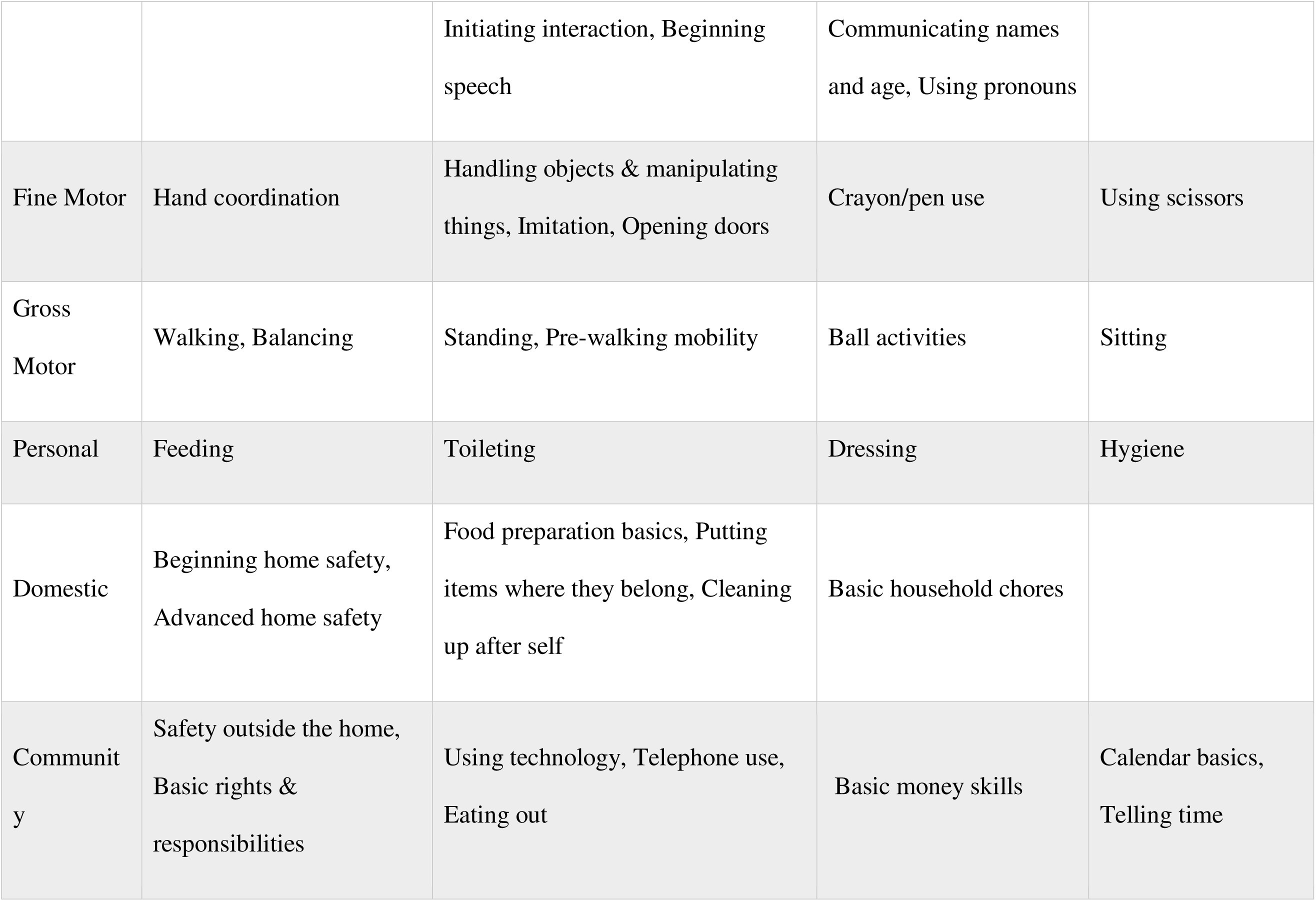
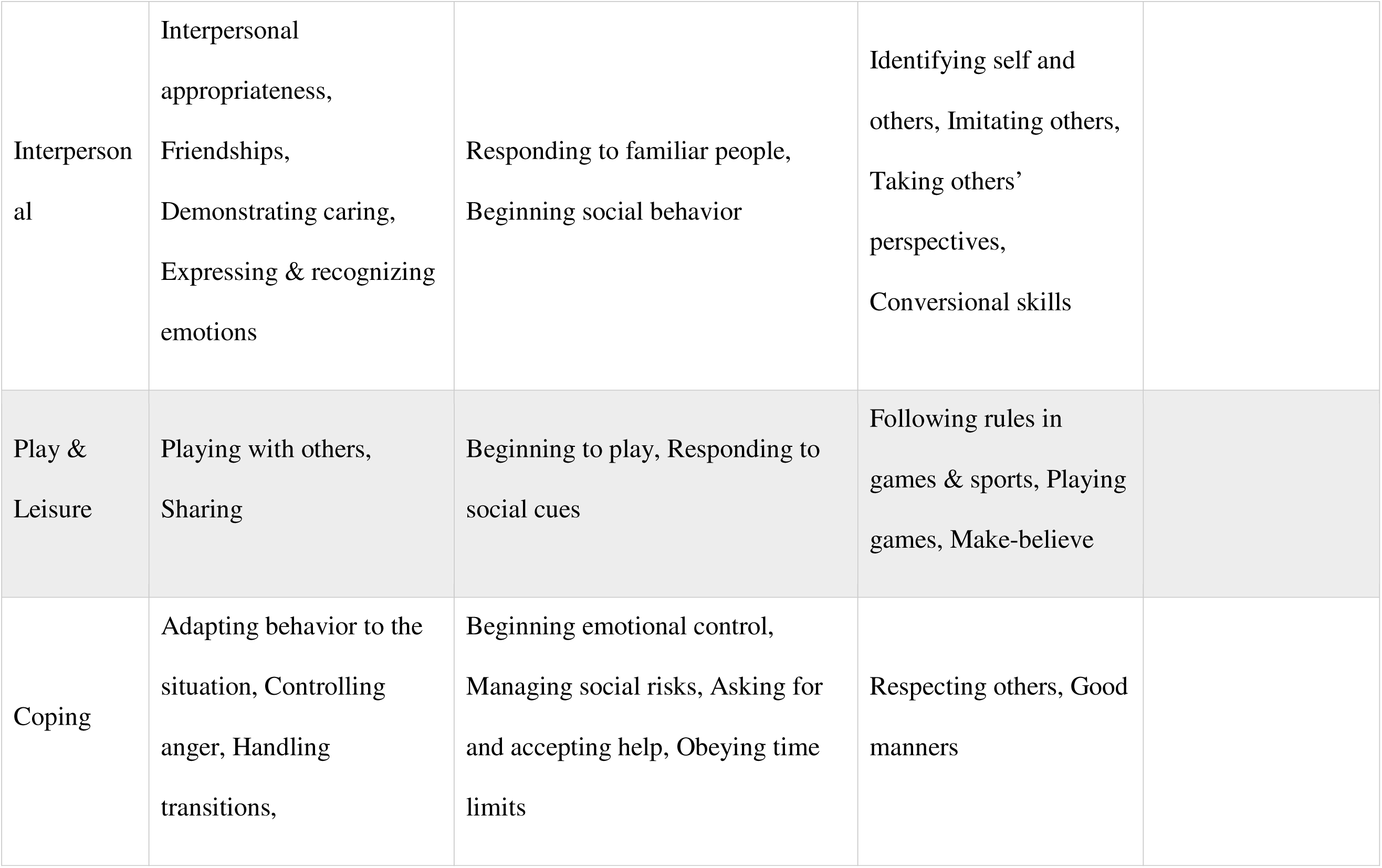
Ratings of the Relevance of Within-Subscale Skills (Clinician Results)

| Subscale | Extremely Important/<br>Highest Importance* | Very Important | Somewhat Important | Not so Important/<br>Least Importance |
| --- | --- | --- | --- | --- |
| Cognitive | Visual attention, Visual spatial skills, Imitation, Memory, Sensorimotor, Problem solving, Object permanence | Understanding pre-academic concepts, Play skills, Visual discrimination |  |  |
| Rec Comm | Understanding words, Following basic instructions, Understanding implied messages | Understanding non-verbal communication, Identifying actions, Attends to entertainment, Responding, Understanding questions, Identifying objects, Following complex instructions | Identifying concepts, Identifying pronouns, Identifying correct syntax |  |
| Exp Comm | Using gestures | Vocabulary, Phrase speech, | Beginning sounds, |  |
|  |  | Initiating interaction, Beginning speech | Communicating names and age, Using pronouns |  |
| Fine Motor | Hand coordination | Handling objects & manipulating things, Imitation, Opening doors | Crayon/pen use | Using scissors |
| Gross Motor | Walking, Balancing | Standing, Pre-walking mobility | Ball activities | Sitting |
| Personal | Feeding | Toileting | Dressing | Hygiene |
| Domestic | Beginning home safety,<br>Advanced home safety | Food preparation basics, Putting items where they belong, Cleaning up after self | Basic household chores |  |
| Community | Safety outside the home,<br>Basic rights & responsibilities | Using technology, Telephone use, Eating out | Basic money skills | Calendar basics,<br>Telling time |

|  |  |  |  |
| --- | --- | --- | --- |
| Interpersonal | Interpersonal appropriateness,<br>Friendships,<br>Demonstrating caring,<br>Expressing & recognizing emotions | Responding to familiar people,<br>Beginning social behavior | Identifying self and others, Imitating others,<br>Taking others' perspectives,<br>Conversational skills |
| Play & Leisure | Playing with others,<br>Sharing | Beginning to play, Responding to social cues | Following rules in games & sports, Playing games, Make-believe |
| Coping | Adapting behavior to the situation, Controlling anger, Handling transitions, | Beginning emotional control,<br>Managing social risks, Asking for and accepting help, Obeying time limits | Respecting others, Good manners |

## Discussion

This study describes a Delphi panel method, aligned with FDA guidance, for generating preliminary data on patient informed MSDs. Consensus ratings of MSDs for Bayley-4 and Vineland-3 subscale GSVs based on caregivers of individuals with AS converged over three iterations with a high degree of agreement. Complete consensus was established for approximately half of all vignettes (100% agreement for 22 of 43 vignettes), while at least 70% agreement was reached for 72% of vignettes. Independent Delphi panels of clinicians who are AS experts and key opinion leaders provided convergent MSD ratings, underscoring the robustness of the MSD ratings generated via caregiver input. Panelists themselves noted that what is considered meaningful is personal and depends on the values of each family, which makes 100% consensus on all subscales unrealistic.

MSD values derived from our study can be used in conjunction with traditional outcome metrics in clinical trials to help interpret results and contextualize findings. This is particularly crucial for therapeutic trials of disease-modifying drugs, which potentially could impact any number of observable outcomes. In conditions like AS, there are often severe, multi-systemic impacts where even small changes in development, adaptive functioning, or behavior could lead to significant improvements in quality of life, thereby representing meaningful change to caregivers.

Our results indicated that using a single MSD rating may not be applicable for most subscales, and depending on baseline functional level may potentially either underestimate or overestimate meaningfulness of treatment benefit. Contrary to expectation, MSD values were unrelated to patient age or molecular subtype (DP vs. DN). Two key factors seemed to drive MSD values obtained for each subscale: 1) developmental level at baseline, and 2) certain highly valued skills that corresponded directly with patient independence and safety (see Tables 4 and 5). These findings have important implications for clinical trials. Consistent with the FDA guidance,^28^ sponsors should consider using baseline level-adjusted MSD estimates to accurately reflect clinical meaningfulness of treatment effects in patients.

In keeping with emerging recommendations on the use of standardized developmental tests as endpoints in clinical trials,^27^ we generated MSDs using GSVs. In order to attach meaning to the GSVs, we characterized them in terms of the content (items) of each scale. While this allowed panelists to understand the types of skills that a specific score would signify, it also may have resulted in undue focus on the skills themselves (e.g., walking) rather than the broader construct (e.g., gross motor). Panelists also noted that it was much easier to understand and conceptualize how the measured skills on the Vineland-3 would impact their quality of life than those on the Bayley-4. In particular, it was particularly challenging to understand the relevance of the skills on the Bayley-4, especially on the Cognitive subscale, which measures foundational skills such as object permanence and cause-and-effect through tasks designed for infants and toddlers. Future research should consider how to balance item-level and construct-level considerations in the development of vignettes.

The MSD values obtained from our study should be considered preliminary and should serve as a foundation for future research. Results of our study should also be compared with anchor-based GSVs, once sufficient data from the Clinical Global Impressions - Improvement (CGI-I) scale are available from the ongoing ASNHS. Additional work is also needed to confirm applicability of MSD ratings obtained across other baseline levels not evaluated in the current study.

It is important to consider other limitations of this work. For example, while the sample size of our Delphi panel for each session (*N*=12-19 caregivers) was consistent with typical panel sizes for in-depth qualitative research^34^, and participants were selected to represent diversity in terms of the age of the individual with AS and molecular subtype, the recruitment of a convenience sample of highly committed parents may limit generalizability of our findings to the broader AS population. Furthermore, the relatively small number of vignettes (i.e., levels of baseline function) per subscale was selected to balance the competing demands of panelist burden and generalizability of findings. Ideally, further follow-up with a larger sample of AS caregivers and a larger number of vignettes representing more baseline functional levels should be conducted to verify the generalizability of our findings. The expanded GSV ranges were added to extend the usability of the MSDs obtained in this study, but these ranges were determined using a set of heuristics rather than empirical data; thus, these ranges should be considered provisional and need to be validated with additional research. Finally, caregivers were asked to make judgments about meaningful change independent of the risk of intervention or the rate of change, which may be critical parameters that caregivers consider when deciding whether to enroll their child in a clinical trial.

Future studies should extend the Delphi and other qualitative or mixed-methods approaches to determine whether our study findings can be replicated among patients and caregivers in other clinical populations that use the Bayley-4 and Vineland-3 as COAs. With sufficient studies, it should be possible to determine whether MSDs derived for different levels of baseline functioning for a particular scale are universal or disease-specific.

## Conclusions

As we enter the second decade of the patient-focused drug development era, we present an overview of the Delphi panel process to collect data on what is meaningful to caregivers of individuals with a rare disorder such as AS. The results of this study serve as one source of preliminary data on MSDs, which can be combined with other study specific outcome metrics to build evidence supporting the thresholds for clinical meaningfulness of treatment effects in future trials. We hope that foundational work from our study spurs additional advancements in the development of MSDs and that a growing body of evidence ultimately creates well-validated processes for developing patient/caregiver informed evaluation criteria for clinical trials.

## Data Availability

All data produced in the present study are available upon reasonable request to the authors

## Authorship Confirmation

All authors certify that they meet the ICMJE criteria for authorship.

