## supplementary table for "Developing Meaningful Score Differences for the Bayley-4 and Vineland-3 in Angelman Syndrome using a Delphi Panel"

### Supplementary Materials

Table of Contents

**1.**	**Schedule of Delphi Panel Meetings**	2

**2.**	**Example of MSD Ratings: Personal Domain (Caregiver and Clinician)**	3

**3.**	**Descriptions of Distribution-Based Metrics**	8

**4.**	**Ratings of Domain and Skill Relevance**	10

**5.**	**Extended Panelist Process Evaluation Survey Questions and Results**	12

#### 1. **Schedule of Delphi Panel Meetings**

| **Meeting** | **Domains Rated** | **Rounds** |
| --- | --- | --- |
| 1 | Intro/Training | -- |
| 2 | Personal (V3) | 1-2 |
| 3 | Personal (V3) | 3 |
|  | Gross Motor (V3 & B4) | 1-2 |
| 4 | Gross Motor (V3 & B4) | 3 |
|  | Fine Motor (V3 & B4) | 1-2 |
| 5 | Fine Motor (V3 & B4) | 3 |
|  | Daily Living: Domestic (V3) | 1-2 |
|  | Daily Living: Community (V3) | 1-2 |
| 6 | Daily Living: Domestic (V3) | 3 |
|  | Daily Living: Community (V3) | 3 |
|  | Receptive Communication (V3 & B4) | 1-2 |
| 7 | Receptive Communication (V3 & B4) | 3 |
|  | Expressive Communication (V3 & B4) | 1-2 |
| 8 | Expressive Communication (V3 & B4) | 3 |
|  | Socialization Interpersonal Relationships (V3) | 1-2 |
|  | Socialization Play and Leisure (V3) | 1-2 |
| 9 | Socialization Interpersonal Relationships (V3) | 3 |
|  | Socialization Play and Leisure (V3) | 3 |
|  | Socialization Coping (V3) | 1-2 |
|  | Socialization Cognitive (B4) | 1-2 |
| 10 | Socialization Coping (V3) | 3 |
|  | Cognitive (B4) | 3 |
|  | Closeout/Finale | -- |

B4 = Bayley-4; V3 = Vineland-3.

#### 2. **Example of MSD Ratings: Personal Domain (Caregiver and Clinician)**

Example of MSD Ratings (Caregiver): Vineland-3 Daily Living Skills – Personal Domain

|  |  | **Low Baseline**  **(GSV = 76; CSEM = 4.1)** | | | **Intermediate Baseline**  **(GSV = 88; CSEM = 3.3)** | | | **High Baseline**  **(GSV = 105; CSEM = 2.5)** | | |
| --- | --- | --- | --- | --- | --- | --- | --- | --- | --- | --- |
| **Subtype** | **Participant** | **R1** | **R2** | **R3** | **R1** | **R2** | **R3** | **R1** | **R2** | **R3** |
| **DN<5yrs** | 1 | 14 | 14 | 14 | 8 | 8 | 8 | 11 | 5 | 1 |
|  | 2 | 14 | 14 | 14 | 8 | 8 | 8 | 1 | 1 | 1 |
|  | 3 | 14 | 20 | 14 | 8 | 8 | 8 | 4 | 9 | 1 |
|  | 4 |  |  | 14 |  |  | 8 |  |  | 1 |
|  | **Median** | **14** | **14** | **14** | **8** | **8** | **8** | **4** | **5** | **1** |
| **DN≥5yrs** | 5 |  |  | 14 |  |  | 8 |  |  | 1 |
|  | 6 | 7 | 10 |  | 6 | 8 |  | 1 | 1 |  |
|  | 7 |  |  | 14 |  |  | 8 |  |  | 1 |
|  | 8 | 16 | 14 |  | 8 | 8 |  | 1 | 1 |  |
|  | 9 | 10 | 10 |  | 10 | 10 |  | 11 | 11 |  |
|  | 10 | 20 | 20 | 14 | 10 | 10 | 8 | 9 | 9 | 9 |
|  | **Median** | **13** | **12** | **14** | **9** | **9** | **8** | **5** | **5** | **1** |
| **DP<5yrs** | 11 | 14 | 14 |  | 8 | 8 |  | 7 | 1 |  |
|  | 12 | 14 | 12 | 14 | 8 | 8 | 8 | 4 | 4 | 1 |
|  | 13 | 12 | 14 | 14 | 6 | 6 | 8 | 4 | 4 | 4 |
|  | 14 | 14 | 14 | 12 | 10 | 8 | 8 | 3 | 3 | 1 |
|  | **Median** | **14** | **14** | **14** | **8** | **8** | **8** | **4** | **3.5** | **1** |
| **DP≥5yrs** | 15 | 16 | 14 |  | 8 | 8 |  | 13 | 7 |  |
|  | 16 | 14 | 14 | 14 | 16 | 8 | 8 | 6 | 3 | 4 |
|  | 17 | 20 | 20 | 14 | 14 | 14 | 8 | 10 | 9 | 1 |
|  | 18 | 14 | 14 | 14 | 6 | 6 | 8 | 9 | 4 | 1 |
|  | 19 | 14 | 14 | 14 | 8 | 8 | 8 | 4 | 4 | 1 |
|  | **Median** | **14** | **14** | **14** | **8** | **8** | **8** | **9** | **4** | **1** |
|  | **Overall Median** | **14** | **14** | **14** | **8** | **8** | **8** | **5** | **4** | **1** |

*Notes*: Where cells are empty, that indicates that the rating was not provided by a particular panelist.
GSV = Growth Scale Value (Range = 10–183). CSEM = Conditional Standard Error of Measurement.
R1= Round 1; R2 = Round 2; R3 = Round 3. DN = Deletion Negative; DP = Deletion Positive.

On the next three pages we provide a visualization of the data in this table that illustrates how panelists converged in their MSD ratings across rounds. Visualizations are illustrated for caregiver panelists only. Clinician panelists demonstrated similar convergence across rounds.

Daily Living - Personal Domain: Caregiver Ratings Across Rounds (Low Baseline)

*y-axis = frequency, x-axis = MSD ratings*

| **Round** | **Low** |
| --- | --- |
| 1 | 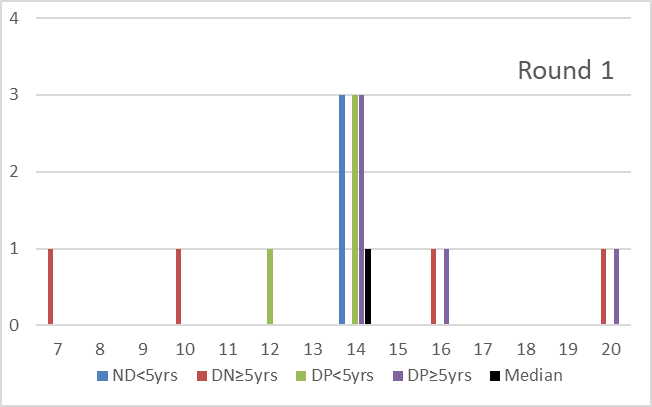 |
| 2 | 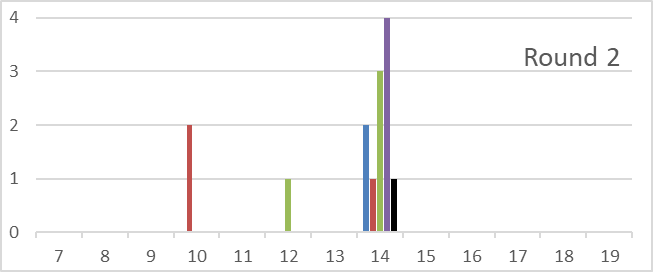 |
| 3 | 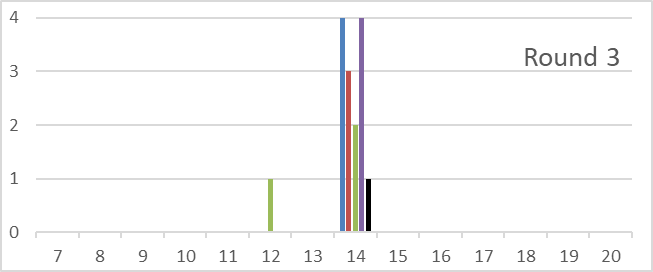 |
|  | Medians Across Rounds: 14 → 14 → 14 |

Daily Living - Personal Domain: Caregiver Ratings Across Rounds (Intermediate Baseline)

*y-axis = frequency, x-axis = MSD ratings*

| **Round** | **Intermediate** |
| --- | --- |
| 1 | 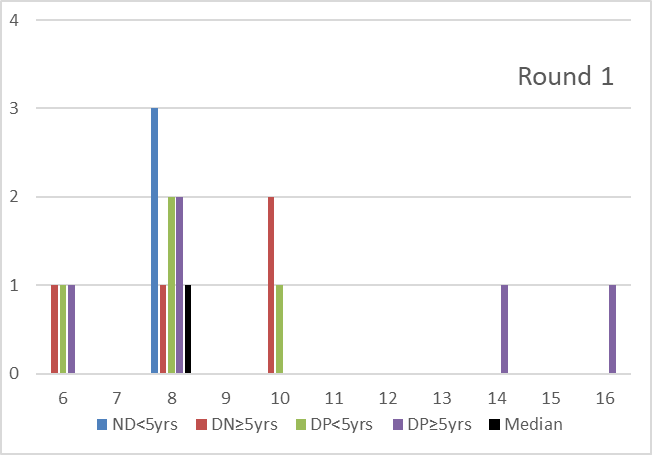 |
| 2 | 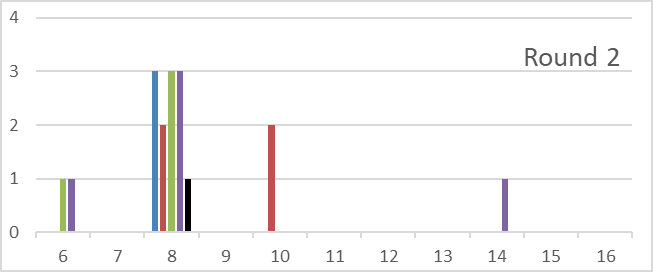 |
| 3 | 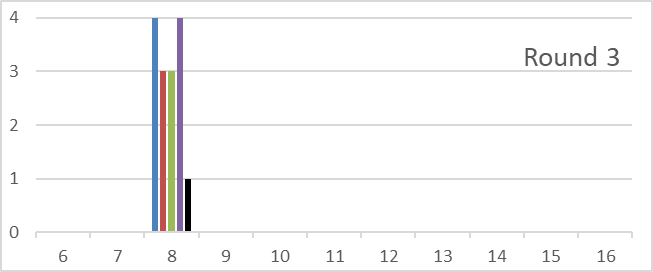 |
|  | Medians Across Rounds: 8 → 8 → 8 |

Daily Living - Personal Domain: Caregiver Ratings Across Rounds (High Baseline)

*y-axis = frequency, x-axis = MSD ratings*

| **Round** | **High** |
| --- | --- |
| 1 | 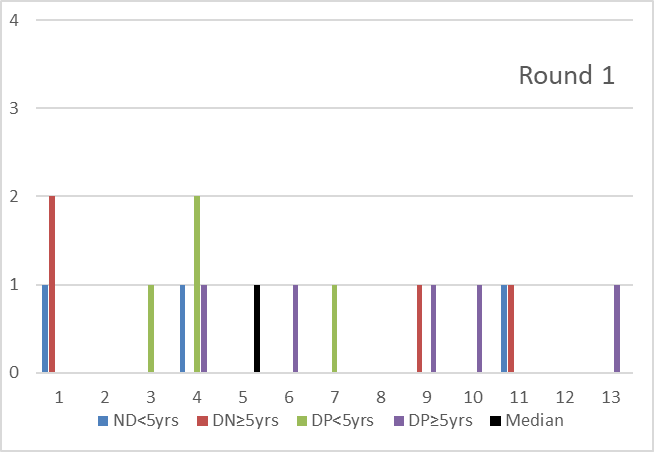 |
| 2 | 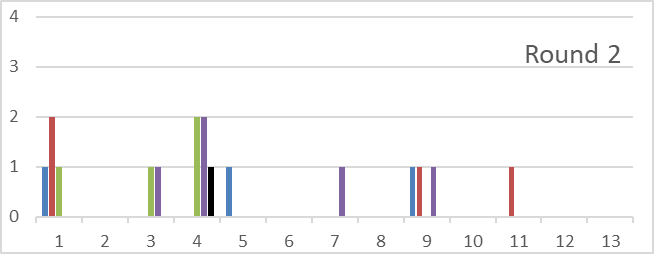 |
| 3 | 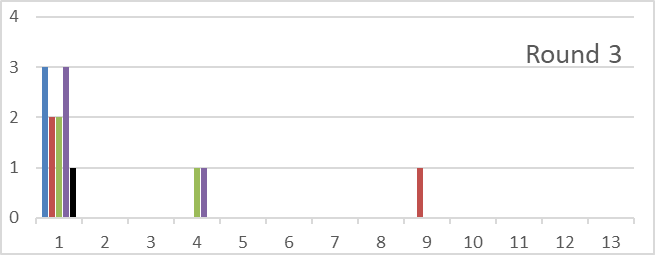 |
|  | Medians Across Rounds: 5 → 4 → 1 |

Example of MSD Ratings (Clinician): Vineland-3 Daily Living Skills – Personal Domain

|  |  | **Low Baseline**  **(GSV = 76; CSEM = 4.1)** | | | **Intermediate Baseline**  **(GSV = 88; CSEM = 3.3)** | | | **High Baseline**  **(GSV = 105; CSEM = 2.5)** | | |
| --- | --- | --- | --- | --- | --- | --- | --- | --- | --- | --- |
| **Subtype** | **Participant** | **R1** | **R2** | **R3** | **R1** | **R2** | **R3** | **R1** | **R2** | **R3** |
| **DN<5yrs** | 1 |  |  | 10 |  |  | 8 |  |  | 1 |
|  | 2 | 10 | 10 | 14 | 6 | 6 | 8 | 5 | 5 | 1 |
|  | 3 | 7 | 7 | 14 | 6 | 6 | 8 | 1 | 1 | 1 |
|  | 4 | 10 | 10 | 14 | 6 | 6 | 8 | 3 | 3 | 1 |
|  | 5 | 10 | 10 | 14 | 2 | 8 | 8 | 3 | 6 | 1 |
|  | **Median** | **10** | **10** | **14** | **6** | **6** | **8** | **3** | **4** | **1** |
| **DN≥5yrs** | 1 |  |  | 10 |  |  | 8 |  |  | 1 |
|  | 2 | 10 | 14 | 14 | 13 | 8 | 8 | 5 | 6 | 1 |
|  | 3 | 14 | 14 | 14 | 8 | 8 | 8 | 1 | 1 | 1 |
|  | 4 | 12 | 12 | 14 | 10 | 10 | 8 | 7 | 7 | 1 |
|  | 5 | 10 | 10 | 14 | 8 | 8 | 8 | 6 | 6 | 1 |
|  | **Median** | **11** | **13** | **14** | **9** | **8** | **8** | **5.5** | **6** | **1** |
| **DP<5yrs** | 1 |  |  | 10 |  |  | 8 |  |  | 1 |
|  | 2 | 7 | 7 | 14 | 2 | 2 | 8 | 1 | 1 | 1 |
|  | 3 | 14 | 10 | 14 | 6 | 2 | 8 | 1 | 1 | 1 |
|  | 4 | 7 | 7 | 14 | 4 | 4 | 8 | 3 | 3 | 1 |
|  | 5 | 10 | 10 | 14 | 8 | 8 | 8 | 6 | 6 | 1 |
|  | **Median** | **8.5** | **8.5** | **14** | **5** | **3** | **8** | **2** | **2** | **1** |
| **DP≥5yrs** | 1 |  |  | 10 |  |  | 8 |  |  | 1 |
|  | 2 | 12 | 12 | 14 | 4 | 4 | 8 | 3 | 3 | 1 |
|  | 3 | 12 | 12 | 14 | 6 | 6 | 8 | 1 | 1 | 1 |
|  | 4 | 12 | 10 | 14 | 6 | 8 | 8 | 1 | 6 | 1 |
|  | 5 | 10 | 10 | 14 | 8 | 8 | 8 | 6 | 6 | 1 |
|  | **Median** | **12** | **11** | **14** | **6** | **7** | **8** | **2** | **4.5** | **1** |
|  | **Overall Median** | **10** | **10** | **14** | **6** | **7** | **8** | **3** | **4** | **1** |

*Notes*: Where cells are empty, that indicates that the rating was not provided by a particular panelist.
GSV = Growth Scale Value (Range = 10–183). CSEM = Conditional Standard Error of Measurement.
R1= Round 1; R2 = Round 2; R3 = Round 3. DN = Deletion Negative; DP = Deletion Positive.

#### 3. **Descriptions of Distribution-Based Metrics**

Distribution-based minimum change thresholds for each of the four AS groups (deletion-positive younger than 5 years old (DP<5yrs), deletion-positive 5 years old or older (DP≥5yrs), deletion-negative younger than 5 (DN<5yrs), deletion-negative 5 years old or older (DN≥5yrs) were computed for each Vineland-3 and Bayley-4 domain. These values were calculated using data from the AS Natural History Study (NHS) and the FREESIAS study. Distribution-based minimum change thresholds included:

1) **The conditional standard error of measurement (CSEM)**. The standard error represents the amount of measurement error involved in a single measurement. These values were provided by the publisher. Values are highest at the extreme ends of the scale and lowest in the middle of the scale. CSEM values were provided in terms of the GSV scale. For measuring change, there are at least two measurements. In this case, the standard error of measurement can be multiplied by 1.96*sqrt(2) = 2.77 to compute the “reliable change index”.^[1]^ The reliable change index is similar to an individual-level test for significance. When a person’s score changes by as much or more than the reliable change index, then that change has a 5% chance or less (i.e., p<0.05) of being observed under the null hypothesis (of no change).
2) **The standard deviation (SD).** Another common distribution-based minimum change threshold is based on the variability in the sample. Commonly, the standard deviation is multiplied by a fractional component (e.g., 0.5, 0.3) to determine a distribution-based change threshold. In this study, we used half a standard deviation (0.5*SD).
3) **The expected 12-month change.** A final distribution-based change threshold was based on observed intra-individual change over time based on NHS data. The number of individuals with more than one score (i.e., longitudinal data) was insufficient for the Bayley-4, but a model-based estimate of 12-month change was computed for all 10 of the Vineland-3 domains. The regression model predicted the average Vineland-3 post-test score (VABS2) using deletion status (DS: deletion positive (1) vs. deletion negative (0)), age category (AGE: less than 5 (0) vs. 5 and greater(1)), number of months between pre-test and post-test (TIME: i.e., 12 months), and Vineland-3 pre-test score (VABS1).

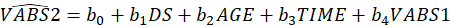

The average pre-test score (
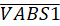
) was then subtracted from the predicted average post-test score (
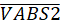
) to obtain the estimated 12-month change score (
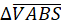
), as illustrated in the equations below for deletion-positive (DP) individuals less than 5 years of age (<5).

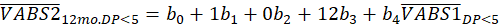

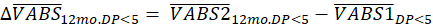

Bayley-4 Distribution-Based Meaningful Score Difference Summary Table

| **MSD Type** | **Subtype** | **Cognitive** | **Rec Comm** | **Exp Comm** | **Fine Motor** | **Gross Motor** |
| --- | --- | --- | --- | --- | --- | --- |
| **CSEM Baseline 1** | **All** | 1.9 | 3.3 | 2.6 | 2.8 | 1.6 |
| **CSEM Baseline 2** | **All** | 1.7 | 2.3 | 2.2 | 2.4 | 2.5 |
| **CSEM Baseline 3** | **All** | 1.5 | 2.4 | 1.9 | 2.4 | ^+^ |
| **CSEM Baseline 4** | **All** | 1.7 | ^+^ | ^+^ | ^+^ | ^+^ |
| **Statistically Significant Difference*** | **All** | 5 | 7 | 7 | 6 | 5 |
| **0.5 SD** | **DN<5yrs** | 3.6 | 3.9 | 4.1 | 3.9 | 7.2 |
|  | **DN**≥**5yrs** | 3.6 | 5.7 | 4.4 | 5.5 | 4.2 |
|  | **DP<5yrs** | 4.3 | 3.8 | 2.4 | 3.8 | 5.3 |
|  | **DP**≥**5yrs** | 5.2 | 5.2 | 3.9 | 3.8 | 3.8 |

*The statistically significant difference is reported in the Bayley-4 manual and is equal to the “reliable change index” described above, that is, a 95% confidence interval around a change score, determined using the standard error of measurement. ^+^This baseline was not included for this subscale. CSEM = conditional standard error of measurement. SD = standard deviation. Rec Comm = Receptive Communication. Exp Comm = Expressive Communication. DN = deletion negative. DP = deletion positive.

Vineland-3 Distribution-Based Meaningful Score Difference Summary Tables

| **MSD Type** | **Subtype** | **Rec Comm** | **Exp Comm** | **Fine Motor** | **Gross Motor** | **Socialization - Coping** |
| --- | --- | --- | --- | --- | --- | --- |
| **CSEM Baseline 1** | **All** | 5.0 | 5.9 | 5.4 | 5.2 | 3.4 |
| **CSEM Baseline 2** | **All** | 3.2 | 4.3 | 4.0 | 3.4 | 2.7 |
| **CSEM Baseline 3** | **All** | 3.1 | 3.5 | 3.8 | 2.9 | 2.6 |
| **Statistically Significant Difference*** | **All** | ~7 | ~8 | ~8 | ~7 | ~6 |
| **0.5 SD** | **DN<5yrs** | 8.2 | 8.9 | 7.1 | 12.8 | 3.7 |
|  | **DN**≥**5yrs** | 7.2 | 12 | 7.8 | 8.6 | 5.4 |
|  | **DP<5yrs** | 8.2 | 7.4 | 9.2 | 14.4 | 3.0 |
|  | **DP**≥**5yrs** | 7.8 | 11 | 6.4 | 9.8 | 4.5 |
| **12 Month Change** | **DN<5yrs** | 4.7 | 12.0 | 5.8 | 11.2 | 2.7 |
|  | **DN**≥**5yrs** | -1.0 | -0.8 | 1.4 | 0.1 | 1.3 |
|  | **DP<5yrs** | 5.7 | 9.4 | 6.0 | 13.0 | 0.0 |
|  | **DP**≥**5yrs** | -0.6 | -1.3 | 0.2 | 0.1 | -0.9 |

*The statistically significant difference is equal to the “reliable change index” described above, that is, a 95% confidence interval around a change score, determined using the standard error of measurement. CSEM = conditional standard error of measurement. SD = standard deviation. Rec Comm = Receptive Communication. Exp Comm = Expressive Communication. DN = deletion negative. DP = deletion positive.

Vineland-3 Distribution-Based Meaningful Score Difference Summary Tables (continued)

| **MSD Type** | **Subtype** | **Socialization - Interpersonal** | **Socialization - Play** | **Daily Living - Personal** | **Daily Living - Domestic** | **Daily Living - Community** |
| --- | --- | --- | --- | --- | --- | --- |
| **CSEM Baseline 1** | **All** | 3.9 | 6.6 | 4.1 | 6.4 | 6.0 |
| **CSEM Baseline 2** | **All** | 3.1 | 3.0 | 3.3 | 4.0 | 3.2 |
| **CSEM Baseline 3** | **All** | 2.6 | 2.6 | 2.5 | ^+^ | ^+^ |
| **Statistically Significant Difference*** | **All** | ~6 | ~6 | ~6 | ~10 | ~10 |
| **0.5 SD** | **DN<5yrs** | 6.6 | 7.8 | 6.1 | 5.6 | 5.2 |
|  | **DN**≥**5yrs** | 6.4 | 7.4 | 7.2 | 7.2 | 6.1 |
|  | **DP<5yrs** | 5.9 | 6.9 | 6.8 | 2.7 | 4.5 |
|  | **DP**≥**5yrs** | 6.0 | 5.9 | 7.0 | 5.7 | 5.8 |
| **12 Month Change** | **DN<5yrs** | 3.7 | 3.2 | 7.7 | 4.5 | 0.4 |
|  | **DN**≥**5yrs** | 1.7 | -0.8 | 1.4 | -2.9 | -2.3 |
|  | **DP<5yrs** | 2.0 | 3.7 | 5.3 | 3.5 | 1.9 |
|  | **DP**≥**5yrs** | -0.1 | -0.4 | -0.3 | -1.4 | -0.6 |

*The statistically significant difference is equal to the “reliable change index” described above, that is, a 95% confidence interval around a change score, determined using the standard error of measurement. ^+^This baseline was not included for this subscale. CSEM = conditional standard error of measurement. SD = standard deviation. DN = deletion negative. DP = deletion positive.

#### 4. **Ratings of Domain and Skill Relevance**

During panel sessions 2–4 we asked participants to rank order skills based on importance. The survey question stated: Please rank these [domain] skills in order of importance (1 being the most important and X being the least important). For sessions 5-10, we changed to a Likert-style scale because some of the domains had many skills and Likert ratings were more efficient than a rank-ordering task. However, this resulted in less comparability in skill importance ratings from the first set of five domains (i.e., Vineland-3 Daily Living Skills Personal, Vineland-3 Gross and Fine Motor, Bayley-4 Gross and Fine Motor), and the other 10 domains. The Likert style survey question participants were asked was: “Please rate the importance of these [domain] skills to your child’s daily functioning or family quality of life”. Response options were on a 5-point Likert scale: 1) Extremely Important, 2) Very Important, 3) Somewhat Important, 4) Not so Important, and 5) Not at all Important. Subskills are defined by the test publisher for Vineland-3. For Bayley-4, the research team created similar subskills for each of the five domains. Subskills by domain are included in the table below.

Domains and Associated Subskills in the AS Vignettes

| **Domain** | **Subskills** |
| --- | --- |
| **Personal (V3)** | Feeding, Dressing, Toileting, Hygiene |
| **Gross Motor**  **(B4 &V3)** | Sitting, Standing, Pre-walking mobility, Walking, Balancing, Ball Activities |
| **Fine Motor**  **(B4 &V3)** | Handling objects and manipulating things, Hand coordination, Crayon/pen use (drawing/coloring), Opening doors, Using scissors, Imitation |
| **Domestic (V3)** | Advanced home safety, Basic household chores, Beginning home safety, Cleaning up after self, Food preparation basics, Putting items where they belong |
| **Community (V3)** | Basic money skills, Basic rights & responsibilities, Safety outside the home, Calendar basics, Eating out, Telephone use, Telling time, Using technology |
| **Receptive Communication**  **(B4 &V3)** | Understanding words, Following basic instructions, Understanding non-verbal communication, Identifying actions, Attending to entertainment, Responding, Understanding questions, Identifying objects, Following complex instructions, Understanding implied messages, Identifying concepts, Identifying pronouns, Identifying correct syntax |
| **Expressive Communication**  **(B4 &V3)** | Using gestures, Vocabulary, Initiating interaction, Using pronouns, Beginning speech, Beginning sounds, Communicating names and age |
| **Interpersonal Relationships (V3)** | Conversional skills, Expressing & recognizing emotions, Friendships, Interpersonal appropriateness, Responding to familiar people, Taking others’ perspectives, Demonstrating caring, Beginning social behavior, Identifying self and others, Imitating others |
| **Play & Leisure (V3)** | Playing with others, Responding to social cues, Beginning to play, Sharing, Playing games, Following rules in games & sports, Make-believe |
| **Coping (V3)** | Controlling anger, Beginning emotional control, Handling transitions, Respecting others, Adapting behavior to the situation, Managing social risks, Asking for and accepting help, Good manners, Obeying time limits |
| **Cognitive (B4)** | Problem solving, Object permanence, Memory, Sensorimotor, Imitation, Visual spatial skills, Understanding pre-academic concepts, Visual attention, Play skills, Visual discrimination |

B4 = Bayley-4; V3 = Vineland-3.

During the final Delphi panel meeting, caregivers were also asked to rate the importance or relevance of the 15 domains in terms of outcomes that matter for individuals with AS and their families. The survey included the question: “Please rate how important it is to include the following domains as endpoints (areas to assess change from the treatment) in future clinical trials”, which had a 4-point Likert scale: 1) Critical to Include, 2) Important to Include, 3) Neutral, and 4) Not Important to Include. Additionally, panelists were asked to select the top 3 domains they felt were most important to include in clinical trials. The results of these questions were summarized in Figure 2 in the body of the manuscript.

#### 5. **Extended Panelist Process Evaluation Survey Questions and Results**

Process Evaluation Survey (Caregiver Results)

| **Meeting 4 Questions (*N*=12):**  Agree/disagree: | | **Percent Agreement** |
| --- | --- | --- |
| - The orientation conducted on July 27, 2022 provided me with a clear understanding of the goals of our project. | | 100% |
| - I have a clear understanding of meaningful change as it is being defined by this project. | | 100% |
| - I am confident about the appropriateness of the final recommendations for meaningful change thresholds for the gross motor domain as discussed earlier in the session today. | | 100% |
| - The facilitators clearly explained the task. | | 100% |
| - The time provided for discussions was adequate. | | 100% |
| - There was an equal opportunity for everyone in my group to contribute his/her ideas and opinions. | | 100% |
| - The discussions after the initial ratings were helpful to me. | | 100% |
| - I was able to follow the instructions and complete the final ratings. | | 100% |
| **Meeting 10 (Finale) Questions (*N*=14):** | |  |
| - Agree/disagree: | |  |
| - I had an opportunity to express my views | | 100% |
| - I found the group discussion to be helpful | | 93%  (1 Neutral) |
| - The facilitators appropriately facilitated the session | | 100% |
| - The definition of meaningful change was easily understood | | 86%  (2 Neutral) |
| - The definition of meaningful change was relevant to me | | 100% |
| - The domains discussed were applicable to my life | | 86%  (1 Neutral,  1 Disagree) |
| - I agree with the final ratings of meaningful change that emerged for the different domains | | 100% |
| - What did you like best about the Meaningful Change Panel meetings? Why? | Caregivers enjoyed hearing the perspectives of other caregivers, enjoyed hearing from very knowledgeable clinicians, and liked the goal of the activity. | |
| - What did you like least about the Meaningful Change Panel meetings? Why? | Timing of meetings and time-consuming nature of meetings. It was sometimes difficult to remember one's ratings and be able to justify them during the meetings. There was some concern that the realities of caregivers with adult AS kids are very different from the realities of caregivers with young AS children. | |
| - What recommendations do you have for the team for ways to improve the process in the future? | Everyone seemed to like the prework (do initial ratings ahead of the meeting, and then come ready to discuss). Need to provide people's ratings back to them so they can come ready to explain their ratings. Need to have more context around the practical implications of different skills before each discussion. One parent indicated concern that consensus wasn't always reached. Should capture more than the rating--also should have the "why" even if caregivers aren't sure. | |
| - Were there any topics or domains that were more difficult to understand or discuss than others? If so, why? What could the team do to make it easier in the future? | Bayley, and especially the cognitive domain was difficult for caregivers to conceptualize. | |
| - What do you think was the most important outcome/result of these panel meetings? | It would be wonderful if researchers understood what is important to AS caregivers and could focus their products or therapies to making life easier for individuals with AS and their caregivers. | |
| - How would you like to see the outcomes of these meetings inform future clinical trials? | Increased focus on measuring things that caregivers value. | |
| - Is there anything you’d like pharmaceutical companies to know as they consider the results of the work we have been doing on meaningful change thresholds? | Safety is one of the most important considerations for caregivers. At this point, kids with AS are vulnerable. Meaningful change would mean any change that results in greater independence and ability to communicate effectively with the outside world. The results of this study came from caregivers who live with their child living with AS every day. Caregivers cannot just pick one area to see improvement in to say whether the drug is working. Important improvement may not be the same for each individual, so it is important to use a variety of domains. | |
| - Is there anything you’d like other parents to know about this work we have been doing on meaningful change thresholds? | Keep expectations high. Meaningful change is different for every individual and family. Thought that the panel did a reasonable job at representing different viewpoints. Hope that the results inform clinical trials. | |
| - Is there anything you’d like to tell the panel investigators now that your participation in this work is complete? | Caregivers found the process meaningful and said it made them hopeful for improvements in treatments and clinical trials. They were thankful to have been a part of the process. | |

*Note*: The last nine questions were open ended. The results are a summary of open-ended responses across all panelists.

Process Evaluation Survey (Clinician Results)

| **Meeting 4 Questions (*N*=4):**  Agree/disagree: | | **Percent Agreement** |
| --- | --- | --- |
| - The orientation conducted on July 27, 2022 provided me with a clear understanding of the goals of our project. | | 100% |
| - I have a clear understanding of meaningful change as it is being defined by this project. | | 100% |
| - I am confident about the appropriateness of the final recommendations for meaningful change thresholds for the gross motor domain as discussed earlier in the session today. | | 100% |
| - The facilitators clearly explained the task. | | 100% |
| - The time provided for discussions was adequate. | | 100% |
| - There was an equal opportunity for everyone in my group to contribute his/her ideas and opinions. | | 100% |
| - The discussions after the initial ratings were helpful to me. | | 100% |
| - I was able to follow the instructions and complete the final ratings. | | 100% |
| **Meeting 10 (Finale) Questions (*N*=4):** | |  |
| - Agree/disagree: | |  |
| - I had an opportunity to express my views | | 100% |
| - I found the group discussion to be helpful | | 100% |
| - The facilitators appropriately facilitated the session | | 100% |
| - The definition of meaningful change was easily understood | | 100% |
| - The definition of meaningful change was relevant to me | | 100% |
| - The domains discussed were applicable to my life | | 75%  (1 Neutral) |
| - I agree with the final ratings of meaningful change that emerged for the different domains | | 100% |
| - What did you like best about the Meaningful Change Panel meetings? Why? | Clinicians enjoyed the format and getting to hear parent perspectives. | |
| - What did you like least about the Meaningful Change Panel meetings? Why? | Process was very time consuming, and not always at an ideal time for everyone's schedule. Bayley items were not very intuitive, making it harder to make judgments. | |
| - What recommendations do you have for the team for ways to improve the process in the future? | Need to provide people's ratings back to them so they can come ready to explain their ratings. Need to have more context around the practical implications of different skills before each discussion. | |
| - Were there any topics or domains that were more difficult to understand or discuss than others? If so, why? What could the team do to make it easier in the future? | Bayley domains were difficult to make meaningful. Should have had videos of the tasks as well as more explanation around how activities extrapolate to real life. | |
| - What do you think was the most important outcome/result of these panel meetings? | Understanding what is important to the families, and informing the FDA about the importance of a multi-domain outcome (one size does not fit all) | |
| - How would you like to see the outcomes of these meetings inform future clinical trials? | Using the values in clinical trials or using the process to change the way clinical trials are evaluated. | |
| - Is there anything you’d like pharmaceutical companies to know as they consider the results of the work we have been doing on meaningful change thresholds? | The results represent the views of a broad group of parents, taking into consideration different ages and subgroups. Even if pharma companies get it, FDA endorsement will be a challenge. Pharma companies need to think about how to incorporate outcomes across multiple domains in clinical trials. | |
| - Is there anything you’d like other parents to know about this work we have been doing on meaningful change thresholds? | [Very little response to this question] | |
| - Is there anything you’d like to tell the panel investigators now that your participation in this work is complete? | Difficult to balance representation (getting all voices represented) with the dynamics of group size. Need to think about how to best incorporate results into clinical trials. | |

*Note*: The last nine questions were open ended. The results are a summary of open-ended responses across all panelists.

1. Jacobson NS, Truax P. Clinical significance: a statistical approach to defining meaningful change in psychotherapy research. *J Consult Clin Psychol*. 1991 Feb;59(1):12-9. doi: 10.1037//0022-006x.59.1.12. PMID: 2002127.
